# Analysis of the Second COVID-19 Wave in India and the United Kingdom Using a Birth-Death Model

**DOI:** 10.1101/2021.06.16.21259009

**Authors:** Narayanan C. Viswanath

**Affiliations:** Government Engineering College, Thrissur, Kerala, India, 680009

**Keywords:** COVID-19 waves, birth-death model, India, the UK

## Abstract

Several countries have witnessed multiple waves of the COVID-19 pandemic between 2020 and 21. The method in [8] is applied here to analyze the COVID-19 waves in India and the UK. For this, a birth-death model is fitted to the active and total cases data for 30 days periods called windows starting from 16^th^ March 2020 up to 10^th^ May 2021. Peculiarities of the parameters suggested a classification of the above windows into three categories: (i) whose fitted parameters predicted a rise in the number of active cases before a fall to zero, (ii) which predicted a decrease, without rising, in the active cases to zero and (iii) which predicted an increase in the active cases until the entire susceptible population gets infected. It follows that some of the type (iii) windows are of the same or lesser concern when compared to some type (i) windows. Further analysis of the type (iii) windows leads to the identification of those which could be indicators of the start of a new wave of the pandemic. The study thus proposes a method for using the present data for identifying pandemic waves in the near future.

## Introduction

[7] applied a generalized birth-death model [2] for modeling the total and actively infected COVID-19 cases in several countries. This model turned out to be a special case of the standard SIR model [3], in which the susceptible cases variable has no explicit role. [8] discussed fitting of the model in [7] when a rise in the number of active cases occurs after a fall. The active and total cases curves of India [9] and the UK [10] are analyzed here to identify the parts of the curve which indicate the occurrence of a new wave. More precisely, the active and total cases curves are observed through 30 days period windows starting from 16^th^ March 2020 up to 10^th^ May 2021. The method in [8] is applied to fit the data observed through a window to a generalized birth-death model [2]. The parameters thus obtained for a window are used to characterize whether it is an indicator of a new wave.

## Methods

By taking the infection birth rate as λ (*t*) = *ae* ^− *bt*^,*a* ≥ 0,*b* ∈ *R* and the recovery rate as μ in a generalized birth-death model [2], [7] derives the following equations for the actively infected *I*(*t*) and total *M*(*t*) cases:

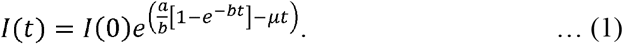

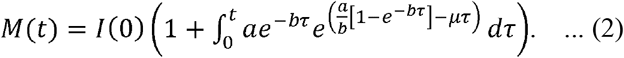

According to [7], a point of inflection on the curve (1) of actively infected cases where its decline starts is given by

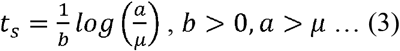

[8] discussed application of the above model when a rise in the number of cases occurs after a fall. There the idea was to fit equations (1) and (2) to a dataset after adjusting the total cases so that the total and active cases remain the same at the starting epoch.

COVID-19 data of active and total cases for India [9] and the UK [10] is analyzed through 30 days period windows starting from 16^th^ March 2020 up to 10^th^ May 2021. For this equations (1) and (2) are fitted to the windowed data, following the method in [8]. Fitting is done by applying the Levenberg-Marquardt nonlinear least-squares algorithm [6] using MATLAB R2019b [5] software.

Depending on the nature of the parameters obtained, each window is classified into one of the following three types:

Type (i): *b*> 0,*a* > µ

In such a period, the infectivity rate λ (*t*) is greater than the recovery rate µ when *t* is close to 0 and becomes less than µ with an increase in time. Consequently, the number of active cases *I*(*t*) first increases in this window attains the peak value at time t_s_, which could be within or outside the window, and then begins to decrease. The number of days from the starting date of a window, after which the number of active cases starts to decline, *Peak_Time* is taken as t_s_, the peak value of active cases, *Active_Peak* is *I*(*t*_*S*_) and the window is called *Rise & Fall*.

Type (ii): *b*> 0,*a* ≤ µ

In such a period, the infectivity rate λ (*t*) is less than the recovery rate µ right from the beginning of the period. Consequently, the number of active cases *I*(*t*) shows a decreasing nature throughout this window. Therefore for such window, *Peak_Time* is taken as 0, *Active_Peak* is taken as *I*(0), and the window is called *Continuously Fall*.

Type (iii): *b*< 0

In such a period, the infectivity rate λ (*t*) is ever-increasing with time. Hence, *I*(*t*) also shows an increasing nature throughout and beyond the window. For such a window, *Peak_Time* could theoretically be the time when every susceptible person at the beginning of the window becomes infected. The number of susceptible cases could be computed using the fitted *I*(*t*) if its value at the beginning of the first window is known. Studies like [4] suggest taking this initial value as the entire population size. However, for a country with a very large population like India, the number of susceptible cases remains close to the initial value. Similar observation can be made for the UK also. In reality, the peak values of *I*(*t*) remain much below the susceptible population size. Hence a theoretical value of *Peak_Time* and *Active_Peak* will be far from reality and may also create panic. Hence these measures will not be defined for the type windows, which will be called *Continuously Rise*.

Since *Peak_Time* is not well defined for the *Continuously Rise* windows, we introduce another measure *Next_Month_New_Cases*, which is defined as follows: Let t_*i* −1_ and t*i*denote the beginning and end of the *i*^th^ window. Let *M*(t_*i* −1_) and *M*(t*i*) denote the fitted total cases for the *i*^th^ window. Let 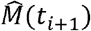 denote the predicted total cases at the end of the (*i+*1)^th^ window, using the parameters for the *i*^th^ window. The predicted total number of new cases during the (*i+*1)^th^ window will then be given by 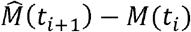. We denote this number by *Next_Month_New_Cases*.

## Results and Discussion

The characterization of 15 windows for India, where the first one ranges from 16^th^ March till 14^th^ April 2020 and the final one ranges from 10^th^ May till 8^th^ June 2021, is given in Figure 1. The parameters for the fit are given in Table 1. Figure 1 identifies the windows starting from the dates 14^th^ June 2020, 13^th^ August 2020, 12^th^ October 2020, 9^th^ February 2021, and 11^th^ March 2021 as those falling in the category of *Continuously Rise*. It follows from Figure 2 that the *Next_Month_New_Cases* value for the 14^th^ June window is close to the 14^th^ July *Rise & Fall* category window, indicating that both windows raise similar concerns. The *Next_Month_New_Cases* value for the *Continuously Rising* 12^th^ October window suggests that this window is of lesser concern when compared to the *Rise & Fall* 12^th^ September window. Thus, the *Continuously Rise* status alone doesn’t characterize a window as a cause for concern. Notice that the huge *Next_Month_New_Cases* value for the continuously rising 13^th^ August window raises the concern that the peak value of active cases may increase. Figure 3 shows that there is an increase in the *Active_Peak* value for the 12^th^ September window, compared to the 14^th^ July window. A reverse nature of the *Peak_Time* for the above windows, as observed in Figure 4, suggests an increase in the active cases in a short period. This can also be observed in Figure 5, which shows the real data of active cases. Hence the concern raised by the 13^th^ August window is worth noting. Thus the *Continuously Rise* status together with the *Next_Month_New_Cases* value characterizes the severity of a window. Notice that the two windows starting from 11^th^ December 2020 and 10^th^ January 2021 suggest a continuous decline in the active cases. After these, the 9^th^ February 2021 window raises concern against a new wave. Notice that this window spans from 9^th^ February to 10^th^ March 2021 and the number of active cases in India was around 200000 in the second week of March 2021, which reached a peak of around 3.7 million in the second week of May 2021 before started declining [9]. Hence a type (iii) window preceded by a type (ii) window needs special attention; especially when it is followed by another type (iii) window. However, these are not the only windows that raise concern. The *Rise & Fall* 15^th^ April 2020 window has a huge *Active_Peak* (please see Figure 3) as well as a long *Peak_Time* (please see Figure 4). Compared to this, the 10^th^ April 2021 window has a short *Peak_Time* but almost double *Active_Peak*. Hence both these windows raise severe concerns.

**Table 1.**
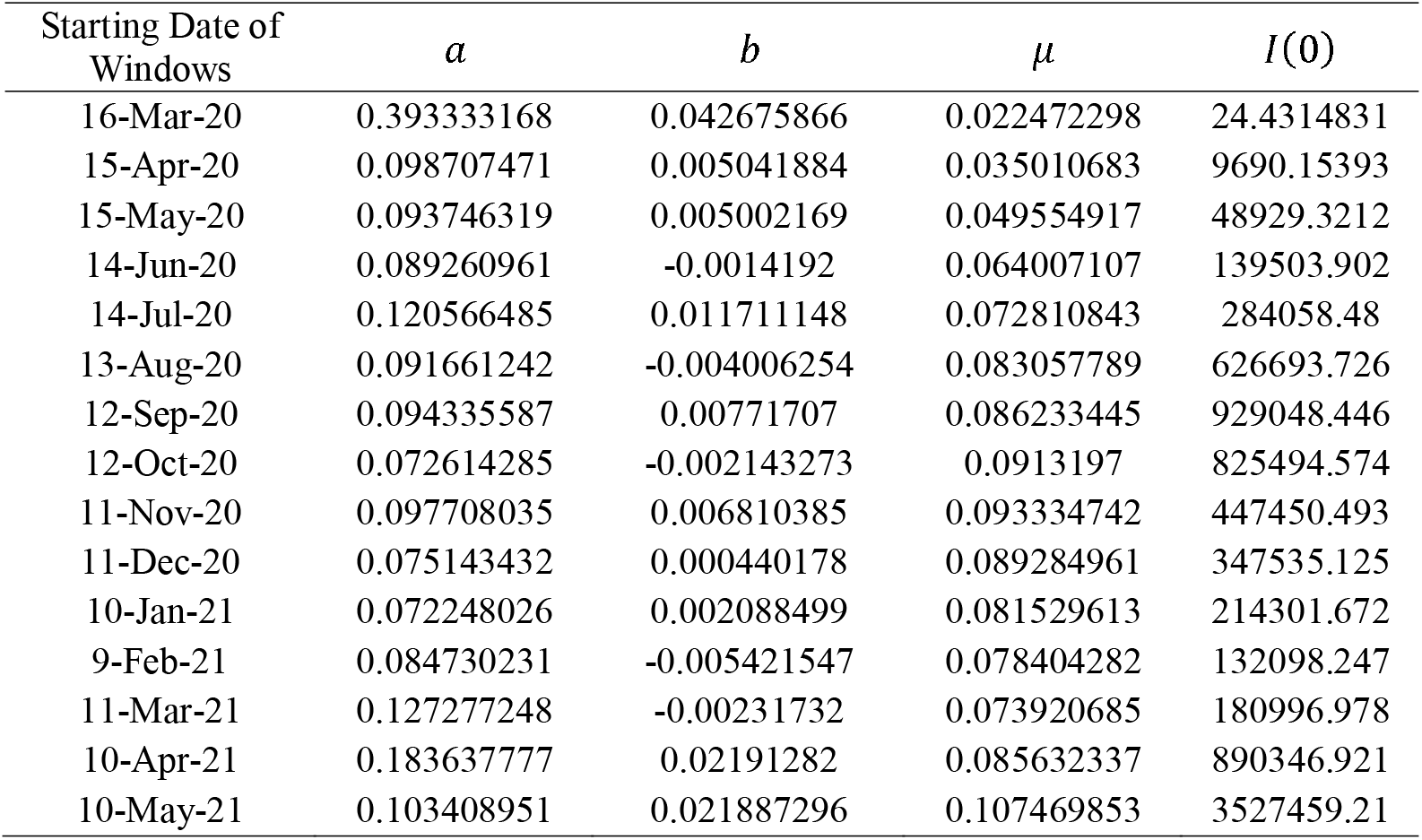
Parameters obtained when models (1) and (2) are fitted for different windowed data of active and total cases for India

| Starting Date of Windows | $a$ | $b$ | $\mu$ | $I(0)$ |
| --- | --- | --- | --- | --- |
| 16-Mar-20 | 0.393333168 | 0.042675866 | 0.022472298 | 24.4314831 |
| 15-Apr-20 | 0.098707471 | 0.005041884 | 0.035010683 | 9690.15393 |
| 15-May-20 | 0.093746319 | 0.005002169 | 0.049554917 | 48929.3212 |
| 14-Jun-20 | 0.089260961 | -0.0014192 | 0.064007107 | 139503.902 |
| 14-Jul-20 | 0.120566485 | 0.011711148 | 0.072810843 | 284058.48 |
| 13-Aug-20 | 0.091661242 | -0.004006254 | 0.083057789 | 626693.726 |
| 12-Sep-20 | 0.094335587 | 0.00771707 | 0.086233445 | 929048.446 |
| 12-Oct-20 | 0.072614285 | -0.002143273 | 0.0913197 | 825494.574 |
| 11-Nov-20 | 0.097708035 | 0.006810385 | 0.093334742 | 447450.493 |
| 11-Dec-20 | 0.075143432 | 0.000440178 | 0.089284961 | 347535.125 |
| 10-Jan-21 | 0.072248026 | 0.002088499 | 0.081529613 | 214301.672 |
| 9-Feb-21 | 0.084730231 | -0.005421547 | 0.078404282 | 132098.247 |
| 11-Mar-21 | 0.127277248 | -0.00231732 | 0.073920685 | 180996.978 |
| 10-Apr-21 | 0.183637777 | 0.02191282 | 0.085632337 | 890346.921 |
| 10-May-21 | 0.103408951 | 0.021887296 | 0.107469853 | 3527459.21 |

**Figure 1.**
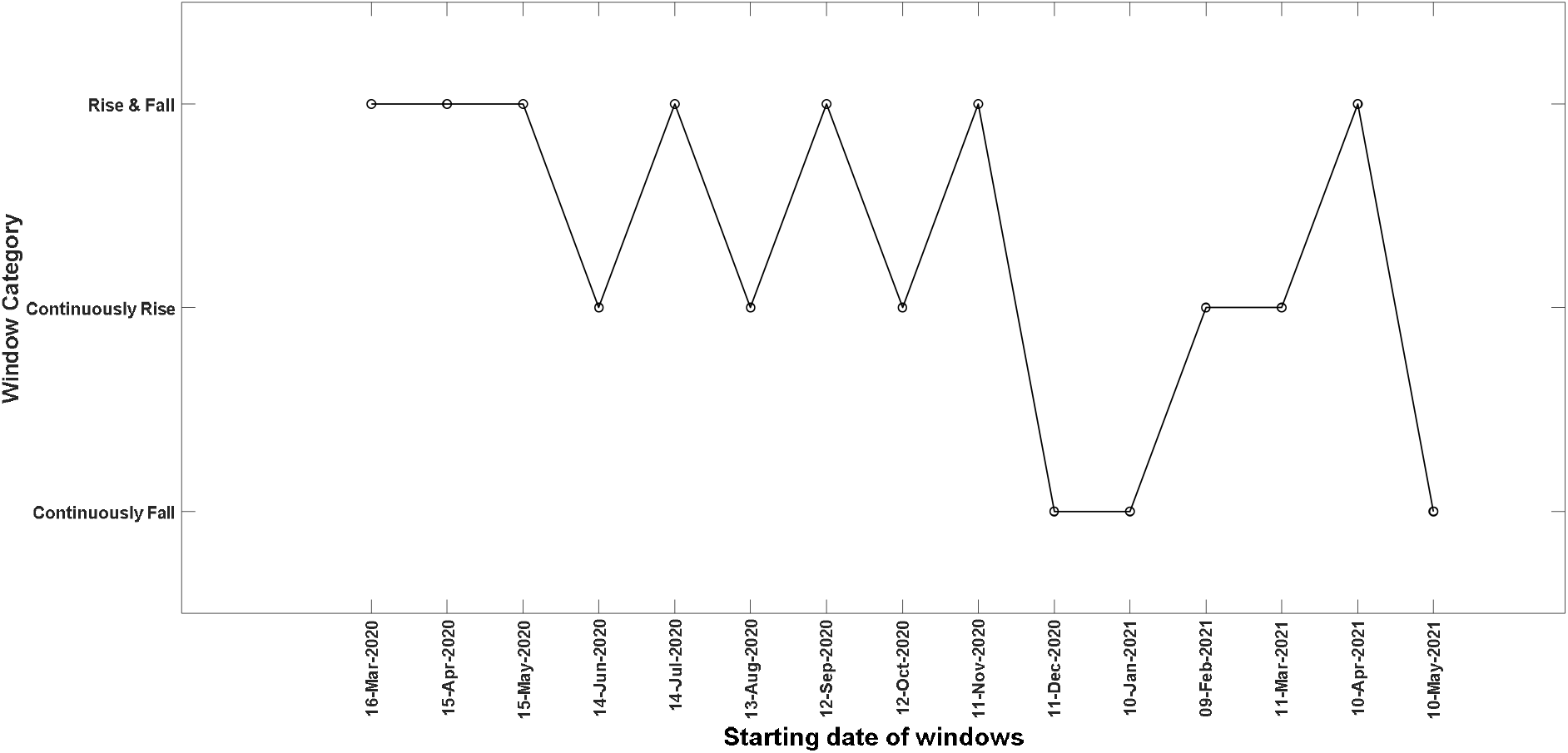
Characteristics of windows starting from 16^th^ March 2020 till 10^th^ May 2021 for India.

**Figure 2.**
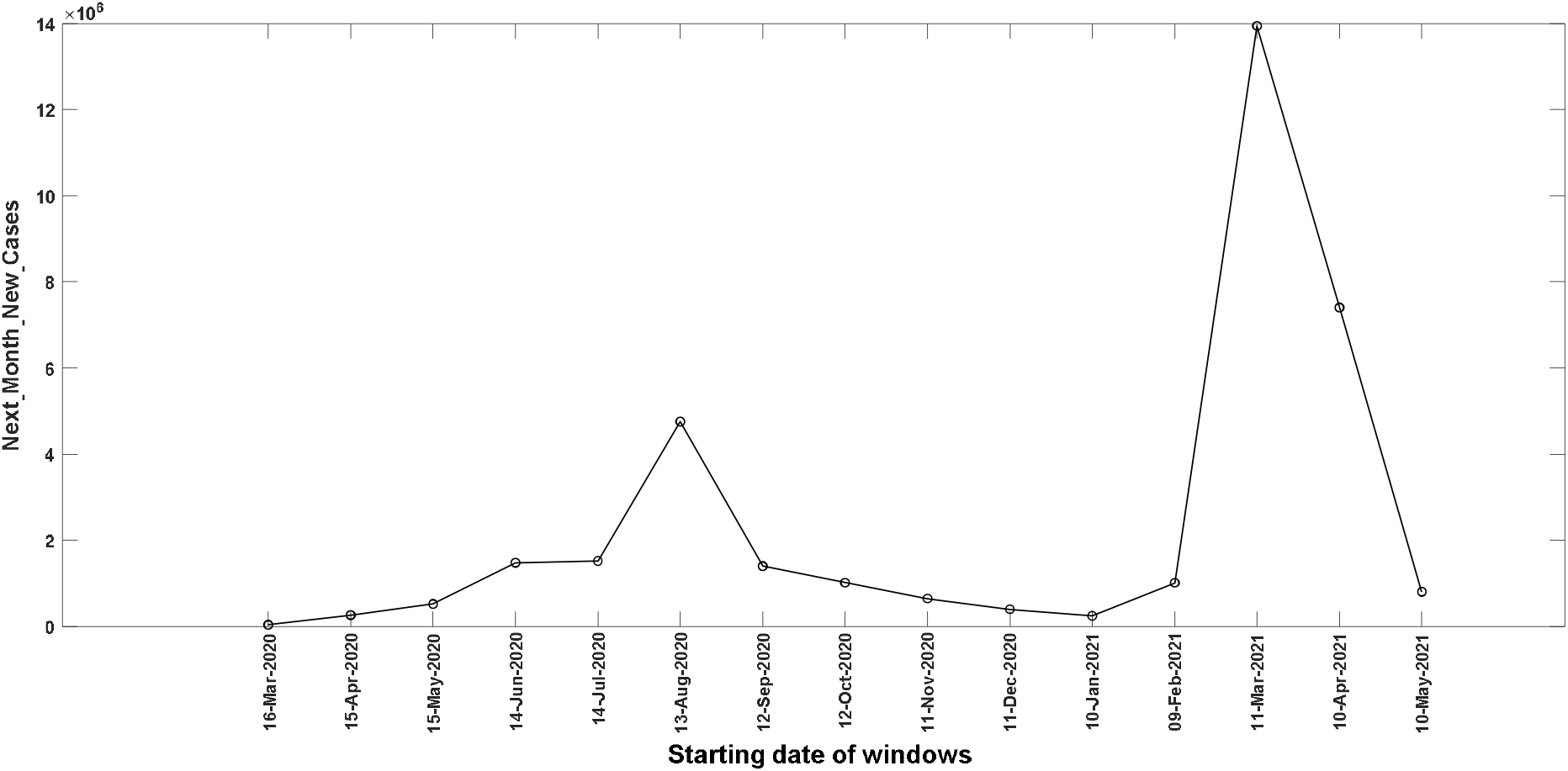
*Next_Month_New_Cases* values for windows from 16^th^ March 2020 till 10^th^ May 2021 for India.

**Figure 3.**
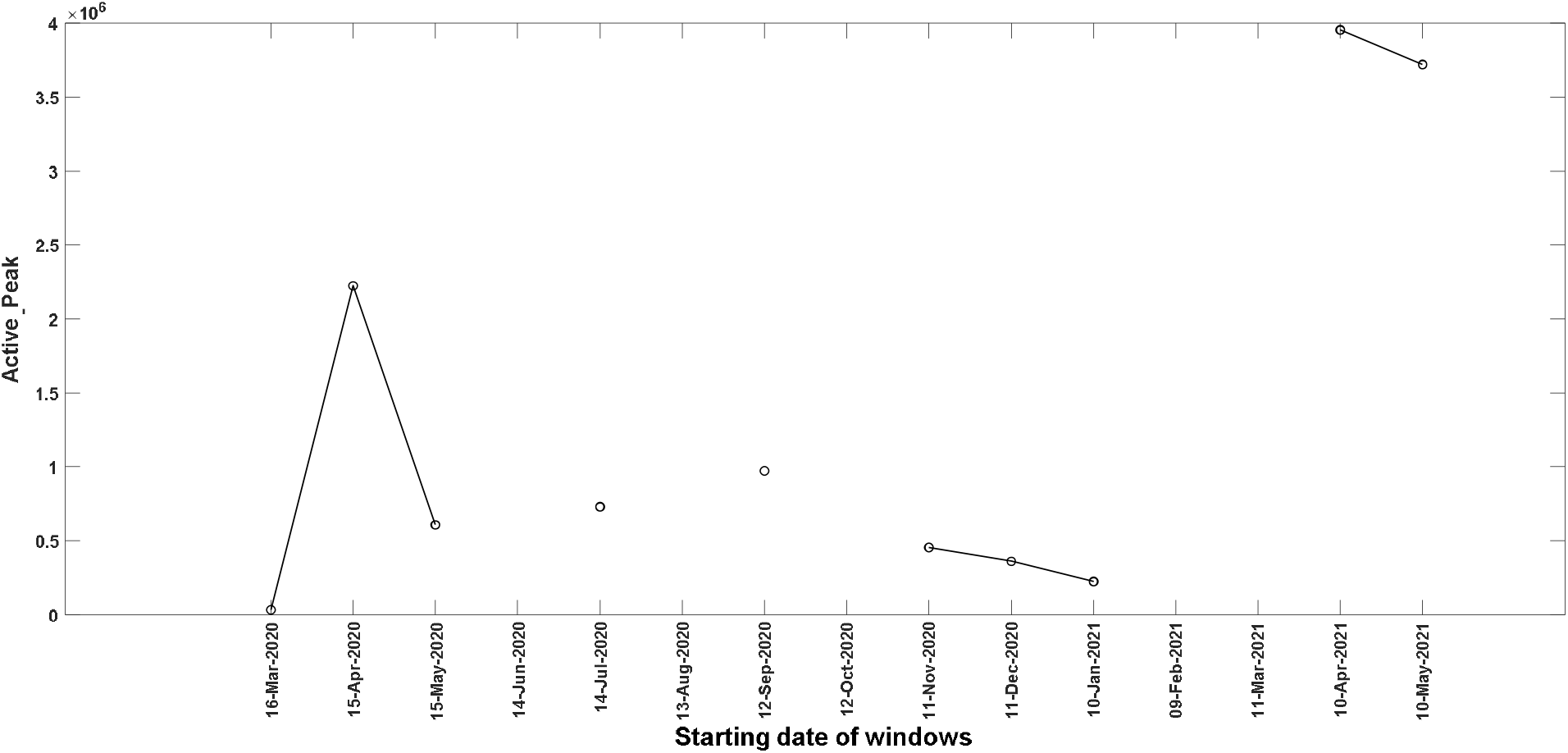
*Active_Peak* values for windows from 16^th^ March 2020 till 10^th^ May 2021 for India.

**Figure 4.**
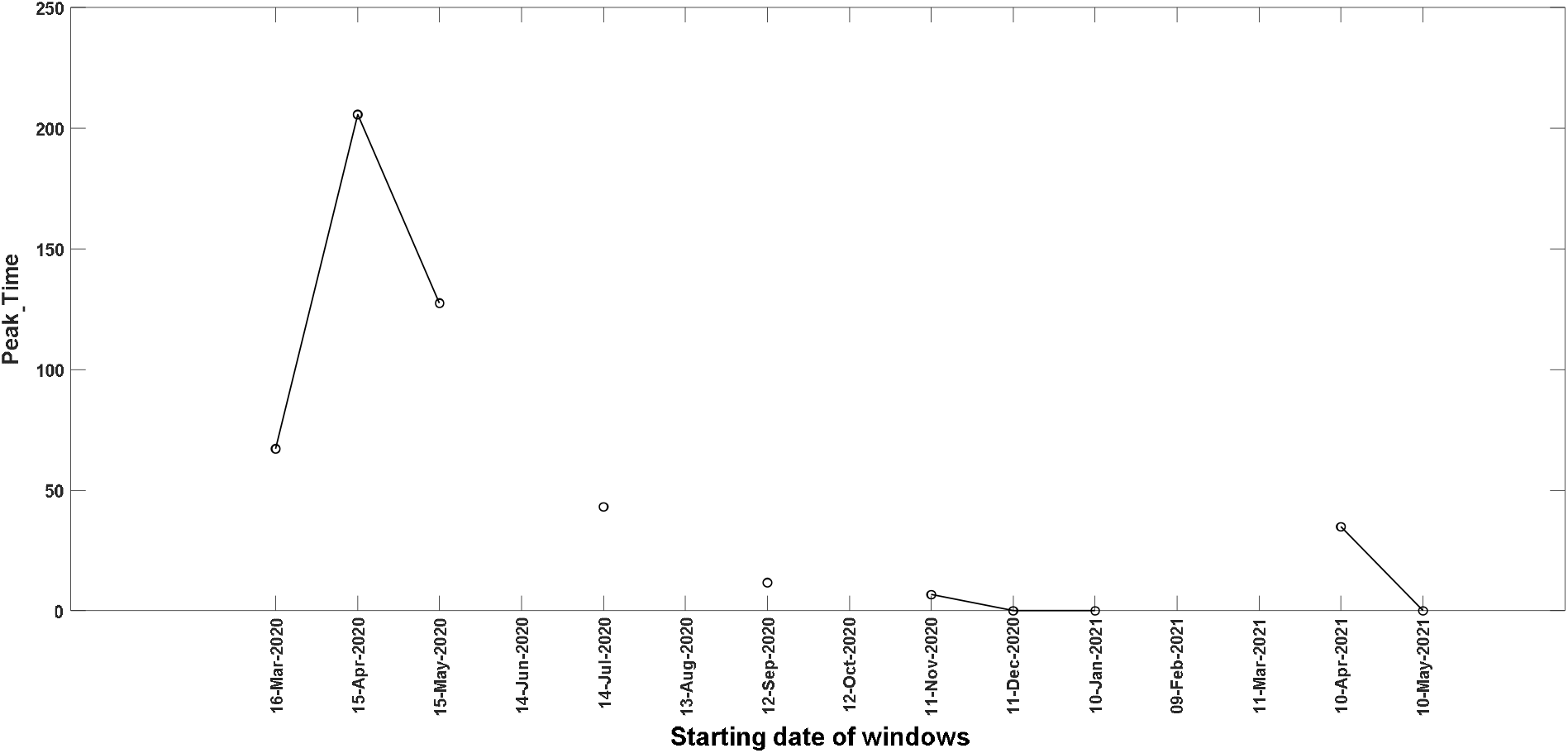
*Peak_Time* values for windows from 16^th^ March 2020 till 10^th^ May 2021 for India.

**Figure 5.**
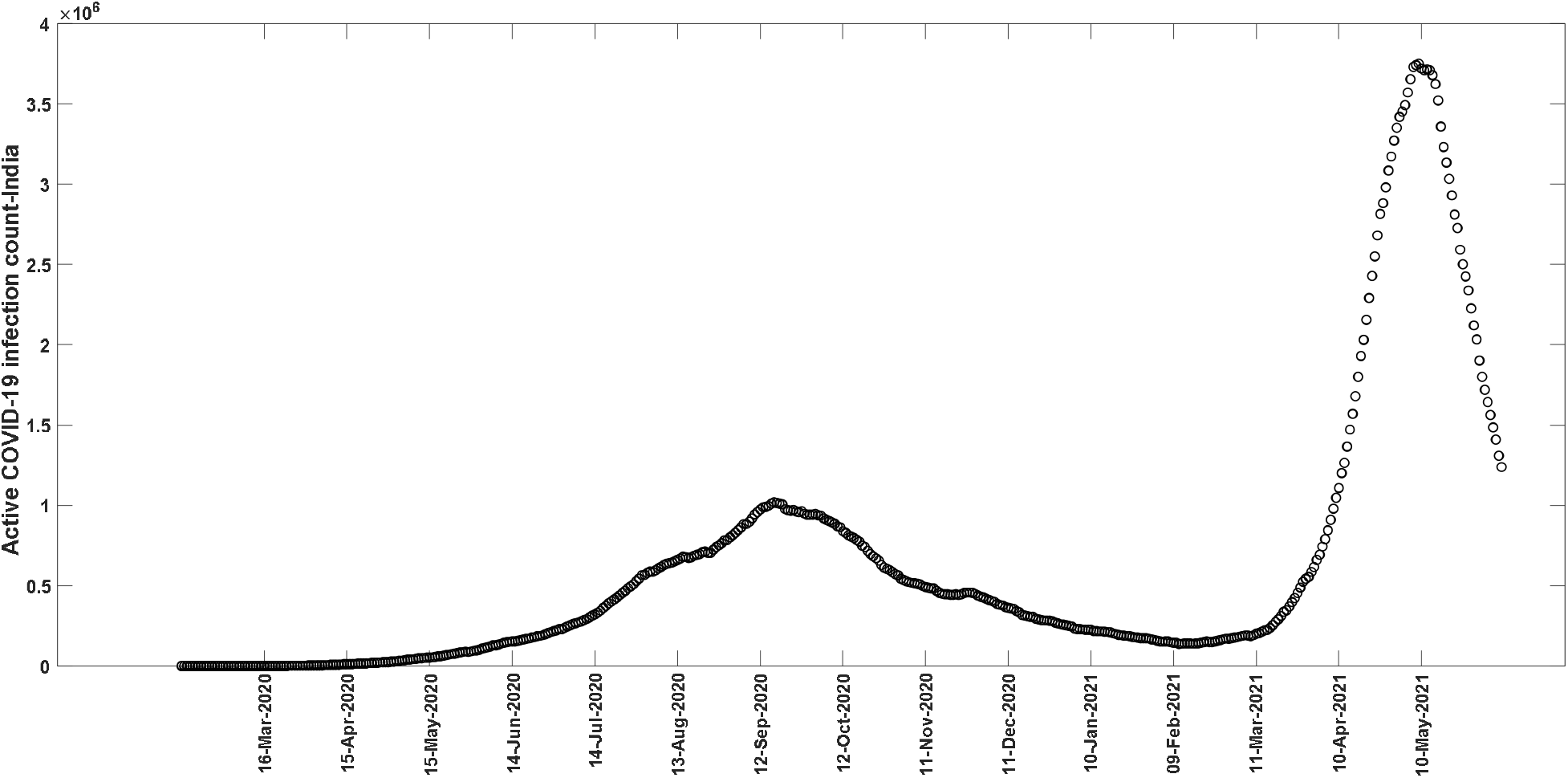
Actual COVID-19 active cases data for India [9].

A comparison of the fitted and actual active and total cases for all the 15 windows for India is given in Appendix I of the supplementary information file (please refer to the odd-numbered figures in there). A comparison of the predicted and actual active and total cases for the nearest future window is also given (please refer to the even-numbered figures in Appendix I of the supplementary information file), where prediction is based on the parameters for the present window. The above figures show that even though the fits are reasonably good, the future predictions give much larger values than the actual counts when the parameters belong to a *Continuously Rise* window. Hence the *Next_Month_New_Cases* measure, which we used to characterize such a window, could be well above the actual count.

The characterization of 15 windows for the UK is given in Figure 6 and the fit parameters are given in Table 2. The *Continuously Rise* windows are those starting from the dates 14^th^ July 2020, 13^th^ August 2020, 12^th^ September 2020, 11^th^ December 2020, 11^th^ March 2021, and 10^th^ May 2021. The 14^th^ July window is preceded by a continuously falling 14^th^ June window. It follows from Figure 7 that the *Next_Month_New_Cases* value for the 14^th^ July window is close to that for the 15^th^ April *Rise & Fall* category window. These together indicate the start of a new wave. This is further strengthened as the 13^th^ August window also belongs to the *Continuously Rise* category and its *Next_Month_New_Cases* value is higher than that for the 14^th^ July window. The actual data in Figure 10 shows that the second wave in the UK got started in September 2020. Hence, in the case of the UK also, a type (iii) window preceded by a type (ii) window needs special attention; especially when it is succeeded by another type (iii) window with a huge *Next_Month_New_Cases* value. Figure 9 shows that there is a sharp decrease in the *Peak_Time* for the *Rise & Fall* 11^th^ November window compared to the 12^th^ October window of the same type. There is also a reduction in the *Active_Peak* value as shown by Figure 8. The above two facts suggest a decline in the active cases. However, the huge *Next_Month_New_Cases* value for the continuously rising 11^th^ December window raises the concern of a rise in the active cases (the concept of a new wave doesn’t apply here as it comes in the middle of a wave). Though the succeeding 10^th^ January and 9^th^ February 2021 windows belong to the *Rise & Fall* category, their huge *Active_Peak* values confirm the concern raised by the 11^th^ December window. The *Next_Month_New_Cases* value for the *Continuously Rise* 11^th^ March 2021 window is similar to that for the same category 14^th^ July 2020 window. Notice that the 14^th^ July window was succeeded by the *Continuously Rise* 13^th^ August 2020 window, which raised the concern of a new wave. Compared to this, the 11^th^ March 2021 window is followed by a *Continuously Fall* 10^th^ April 2021 window, which dismisses the concern of another wave for a short period. However, the *Continuously Rise* status of the 10^th^ May 2021 window together with the huge *Next_Month_New_Cases* value, which is even higher than the 12^th^ September 2020 window renew the concern of a third wave in the UK.

**Table 2.** Parameters obtained when models (1) and (2) are fitted for different windowed data of active and total cases for the UK.

| Starting Date of Windows | $a$ | $b$ | $\mu$ | $I(0)$ |
| --- | --- | --- | --- | --- |
| 16-Mar-20 | 0.324102489 | 0.048528408 | 0.021023342 | 790.180606 |
| 15-Apr-20 | 0.055124626 | 0.030072888 | 0.005794789 | 70904.0454 |
| 15-May-20 | 0.03372626 | 0.121190795 | 0.016458604 | 175230.636 |
| 14-Jun-20 | 0.009442344 | 0.022801929 | 0.03685976 | 130874.459 |
| 14-Jul-20 | 0.010319374 | -0.037222901 | 0.031680145 | 51497.5783 |
| 13-Aug-20 | 0.018519453 | -0.036613757 | 0.015659297 | 36294.6964 |
| 12-Sep-20 | 0.050329258 | -0.009301011 | 0.007419923 | 61138.7187 |
| 12-Oct-20 | 0.062524315 | 0.028824707 | 0.004455551 | 270926.731 |
| 11-Nov-20 | 0.033176081 | 0.048165502 | 0.011380747 | 832368.307 |
| 11-Dec-20 | 0.023246957 | -0.027147445 | 0.020268453 | 957667.528 |
| 10-Jan-21 | 0.035225491 | 0.055663458 | 0.009290769 | 1529112.68 |
| 9-Feb-21 | 0.033014621 | 0.212817632 | 0.030173346 | 1805801.67 |
| 11-Mar-21 | 0.004710081 | -0.043640352 | 0.056521665 | 744606.225 |
| 10-Apr-21 | 0.016413397 | 0.010512717 | 0.034801206 | 189597.78 |
| 10-May-21 | 0.016580241 | -0.04346663 | 0.027449732 | 96862.1512 |

**Figure 6.**
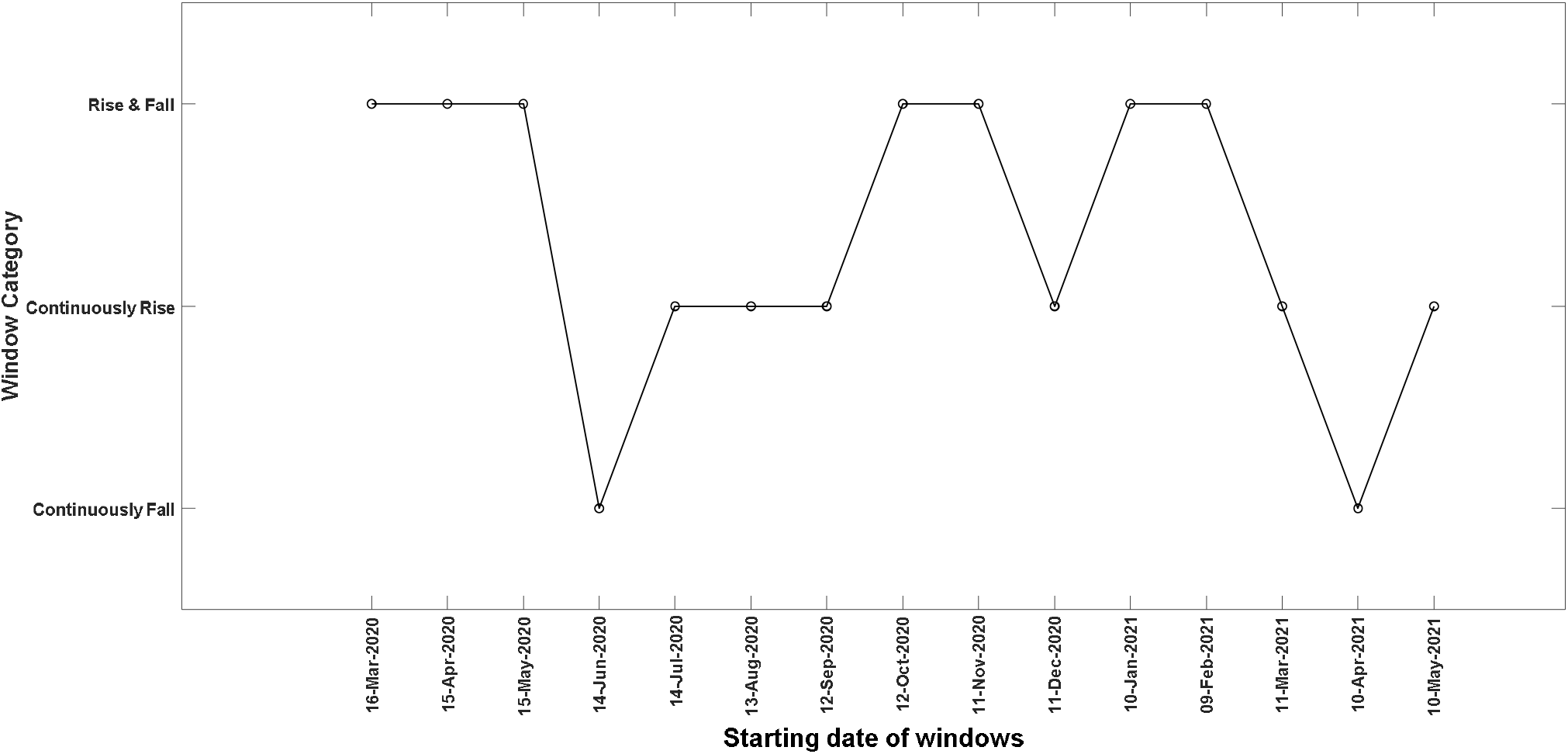
Characteristics of windows starting from 16^th^ March 2020 till 10^th^ May 2021 for the UK.

**Figure 7.**
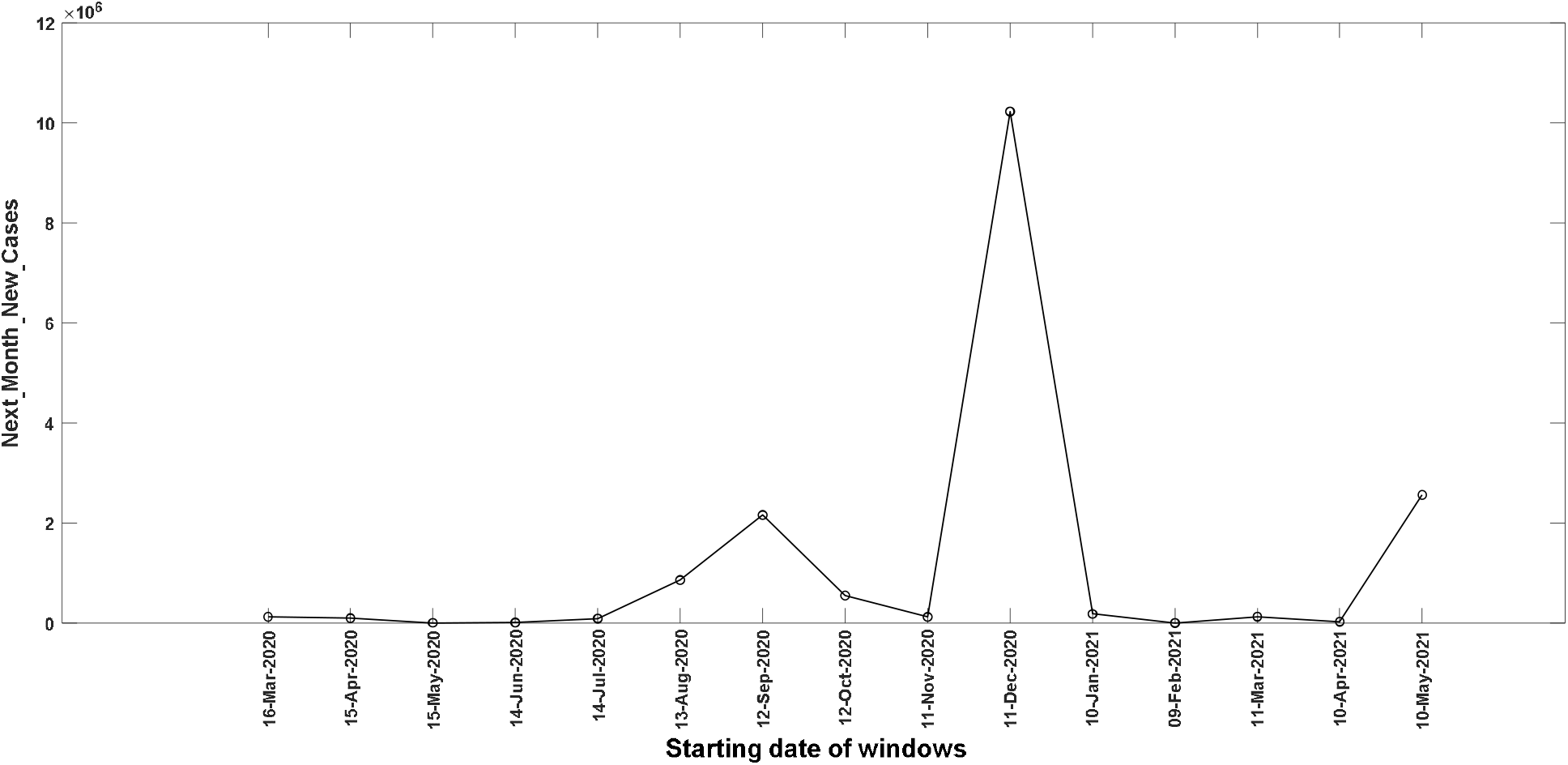
*Next_Month_New_Cases* values for windows from 16^th^ March 2020 till 10^th^ May 2021 for the UK.

**Figure 8.**
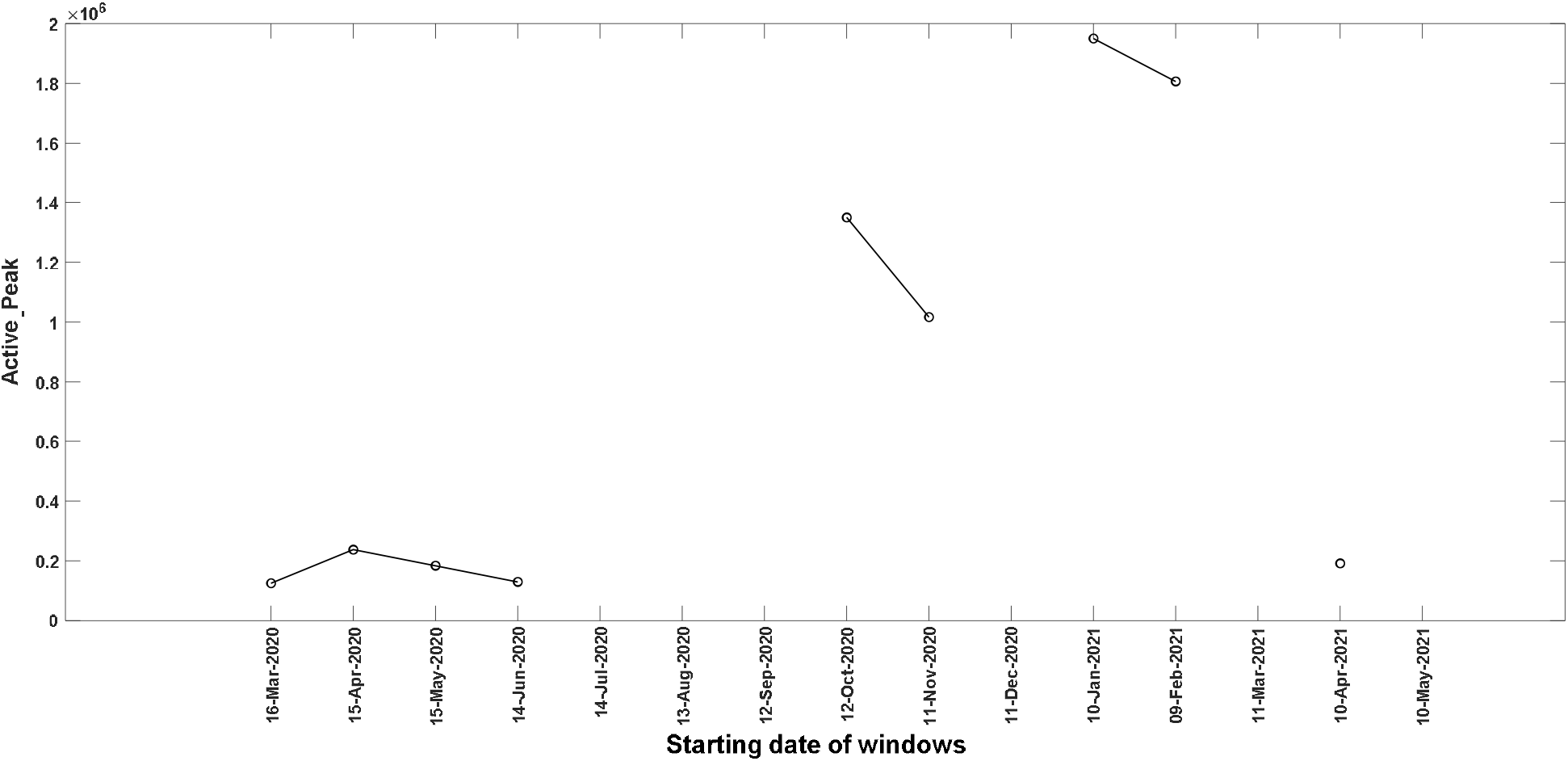
*Active_Peak* values for windows from 16^th^ March 2020 till 10^th^ May 2021 for the UK.

**Figure 9.**
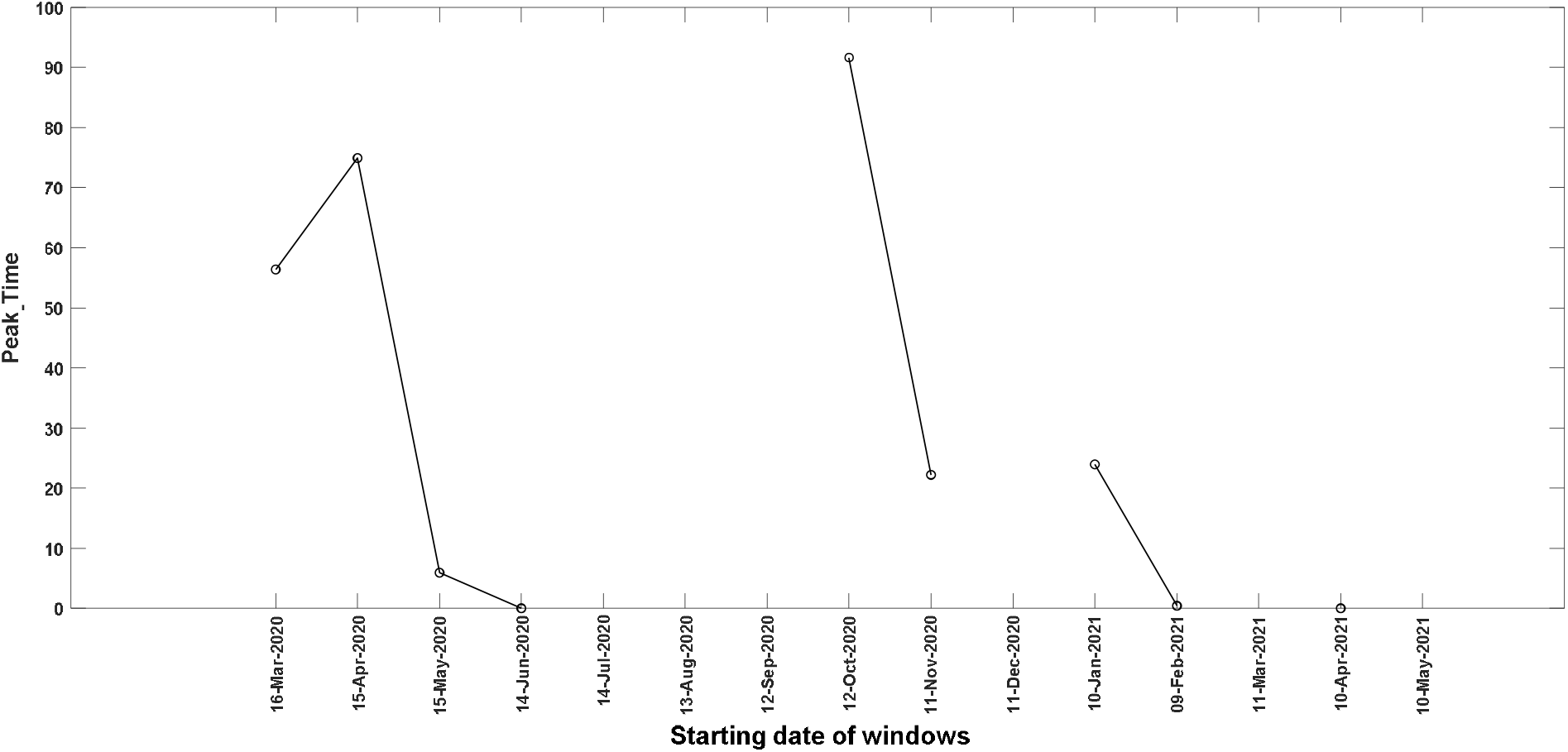
*Peak_Time* values for windows from 16^th^ March 2020 till 10^th^ May 2021 for the UK.

**Figure 10.**
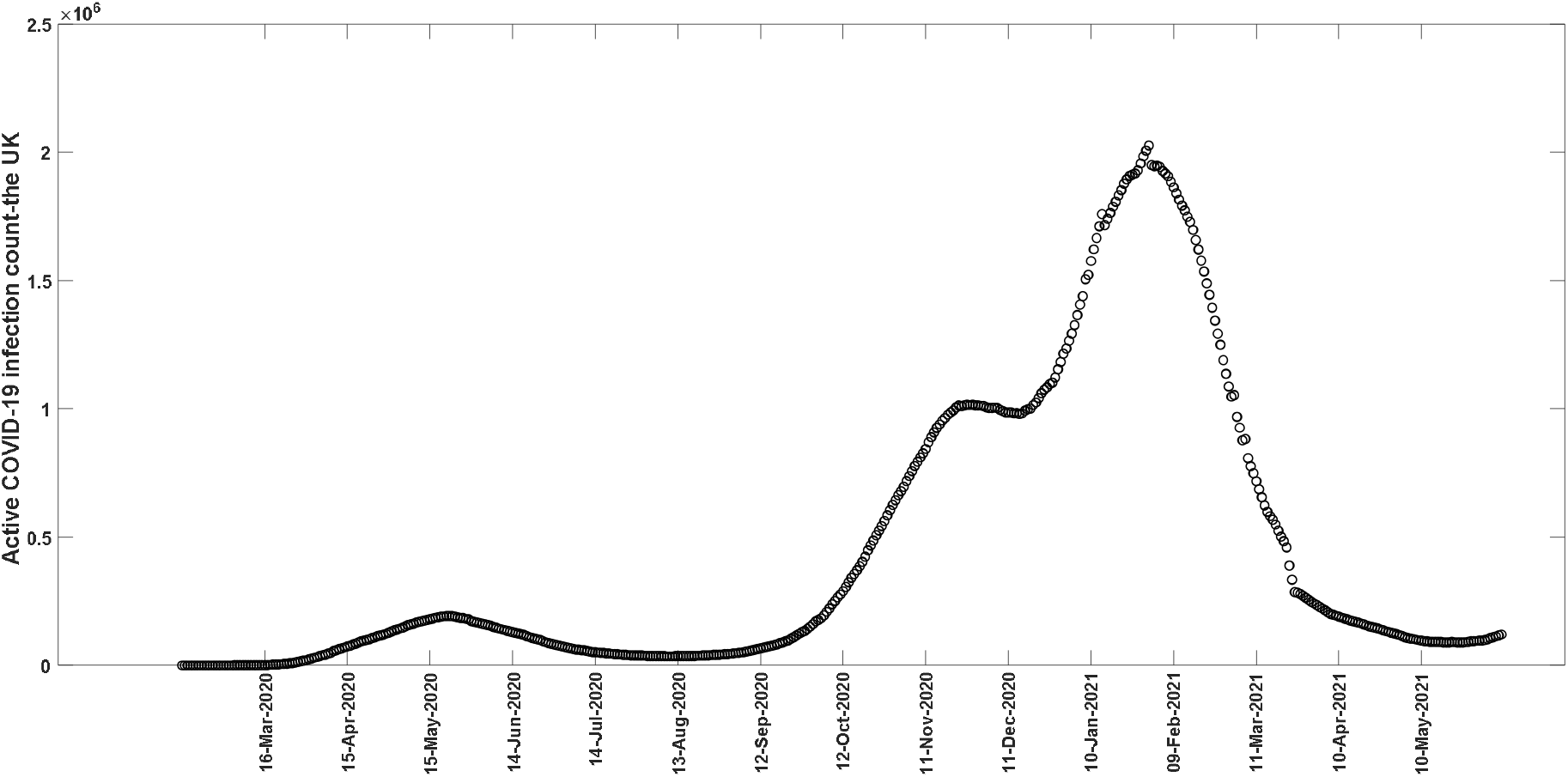
Actual COVID-19 active cases data for the UK [10].

A comparison of the fitted and the actual active and total cases for all the 15 windows for the UK is given in Appendix II of the supplementary information file (please refer to the odd-numbered figures in there). A comparison of the predicted and actual active and total cases for the nearest future window is also given (please refer to the even-numbered figures in Appendix II of the supplementary information file), where prediction is based on the parameters for the present window. The above figures show that in the case of the UK also, the fits are reasonably good. However, the future predictions are worse than in the case of India in the sense that much larger values are obtained than the actual counts, especially when the parameters belong to a *Continuously Rise* window. Also, notice that the current model doesn’t consider the effect of the ongoing vaccination program and other containment measures. Therefore the indication of the third wave by the 10^th^ May 2021 window needs further examination in light of the above facts. This is important since the epidemic forecasting of the COVID-19 pandemic had several setbacks [1].

## Conclusions

The study proposes a method of windowing the data of active and total COVID-19 cases for using the present data for identifying pandemic waves in the near future. 30 days period windows are used here, where the first one ranges from 16^th^ March till 14^th^ April 2020, and the final one ranges from 10^th^ May till 8^th^ June 2021. Depending on the nature of their parameters, the windows are classified into three categories: *Rise & Fall, Continuously Fall*, and *Continuously Rise*. A *Continuously Rise* window is analyzed in light of the *Next_Month_New_Cases* value to determine whether it indicates a new wave when preceded by a *Continuously Fall* window. *Continuously Rise* windows are identified, which recommended the second wave in India and the UK. In the case of the UK, the 10^th^ May 2021 window is indicating the third wave. However, the differences between the future predictions and actual values and the fact that the model doesn’t consider the effect of vaccination and other COVID-19 containment measures demand further investigation of the above prediction.

The current study starts windowing from 16^th^ March 2020 and uses 30 days windows. Studies on the effect of the initial date and the window length on the predictions and analysis seem interesting. We wish to do this in future studies.

## Supporting information

Comparison of the fitted/predicted values with the actual counts

## Data Availability

The data are available from https://www.worldometers.info/coronavirus/country/india
https://www.worldometers.info/coronavirus/country/uk

https://www.worldometers.info/coronavirus/country/uk

