## Supplementary material for "Analysis of the Second COVID-19 Wave in India and the United Kingdom Using a Birth-Death Model": Comparison of the fitted/predicted values with the actual counts

**Appendix I**

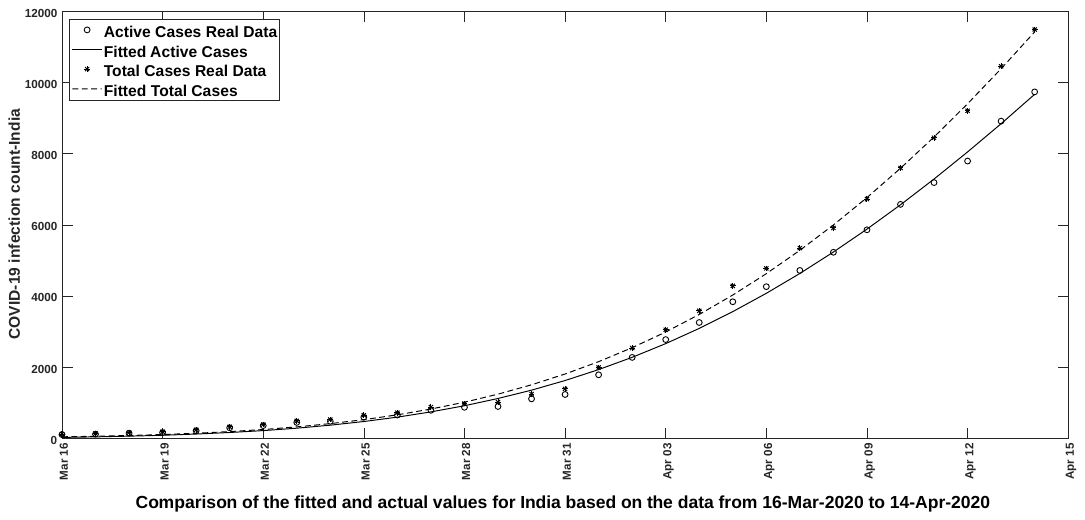

Figure 1. Comparison of the fitted and actual values for India based on the window starting

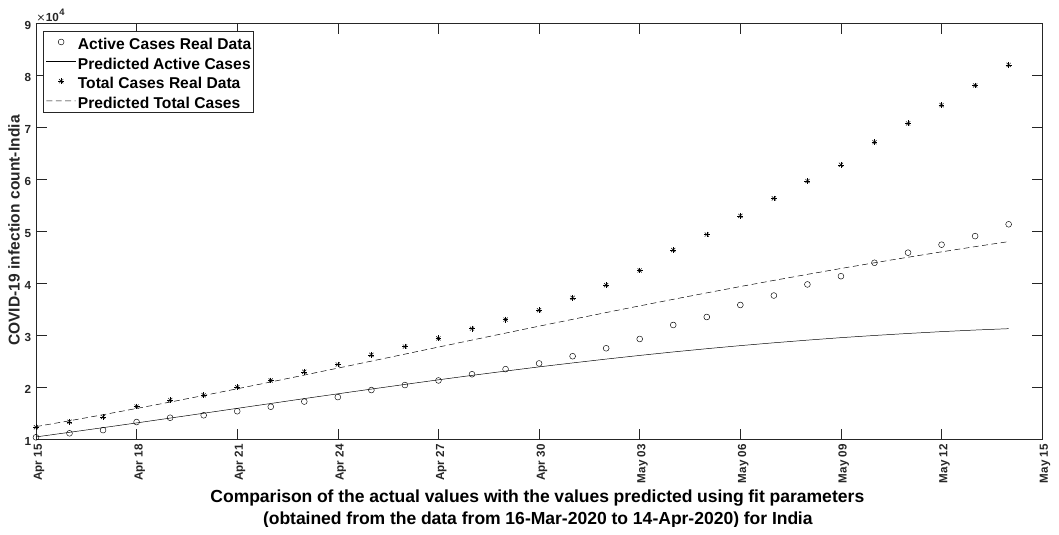

**Figure 2**

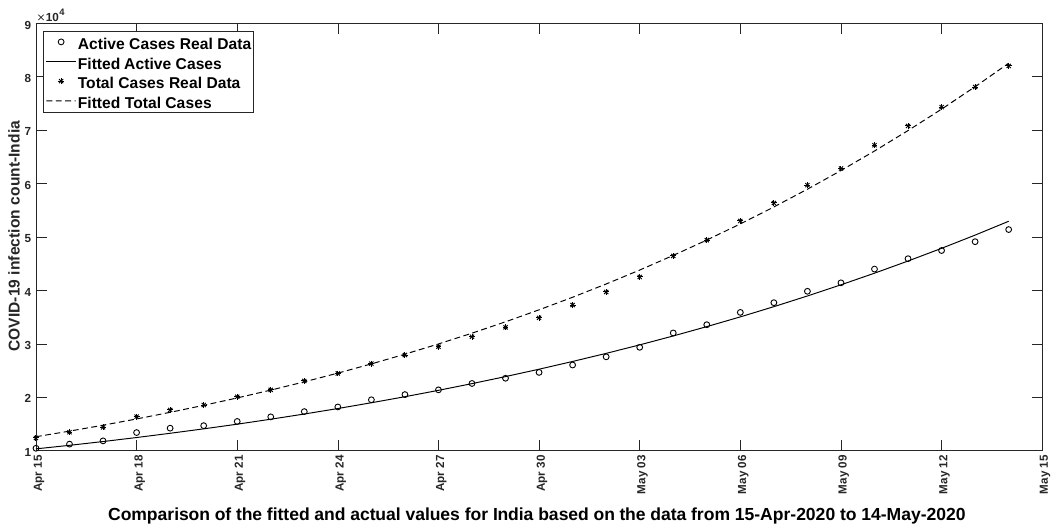

**Figure 3**

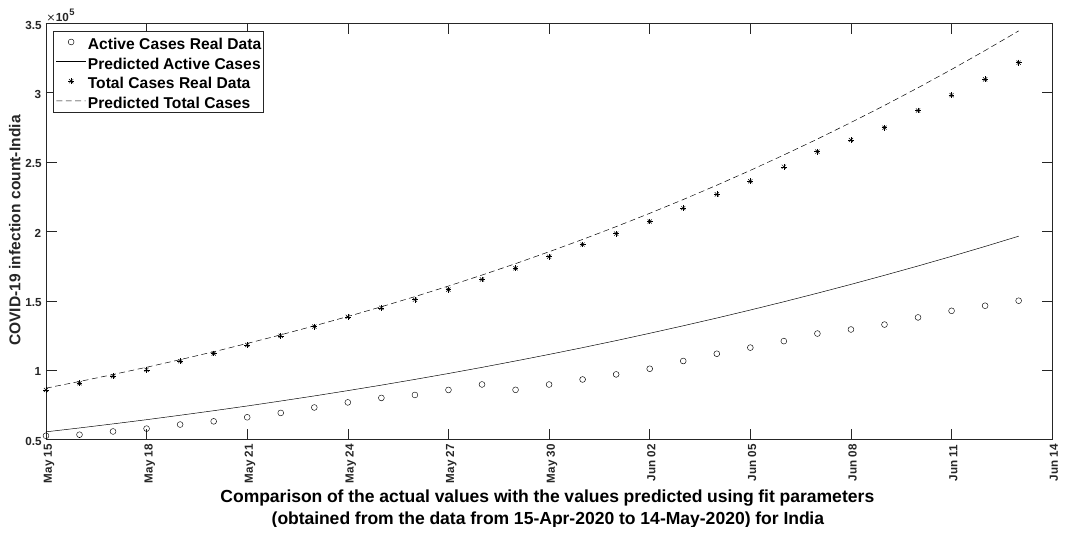

**Figure 4**

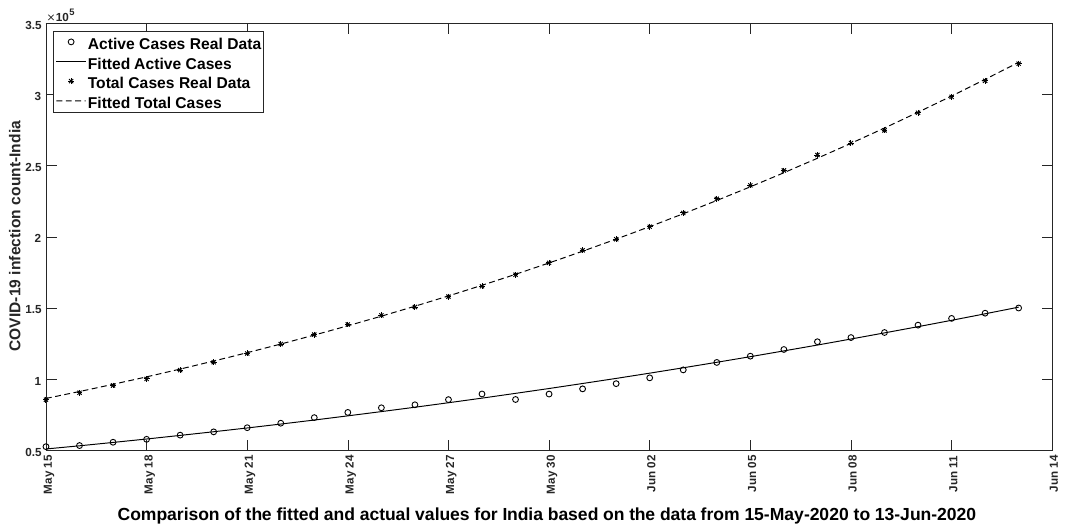

**Figure 5**

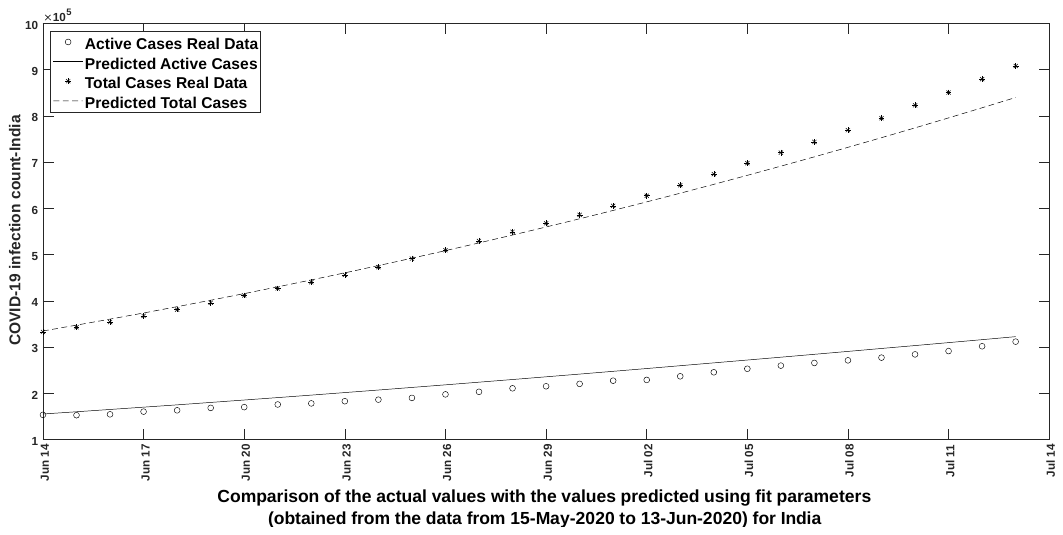

**Figure 6**

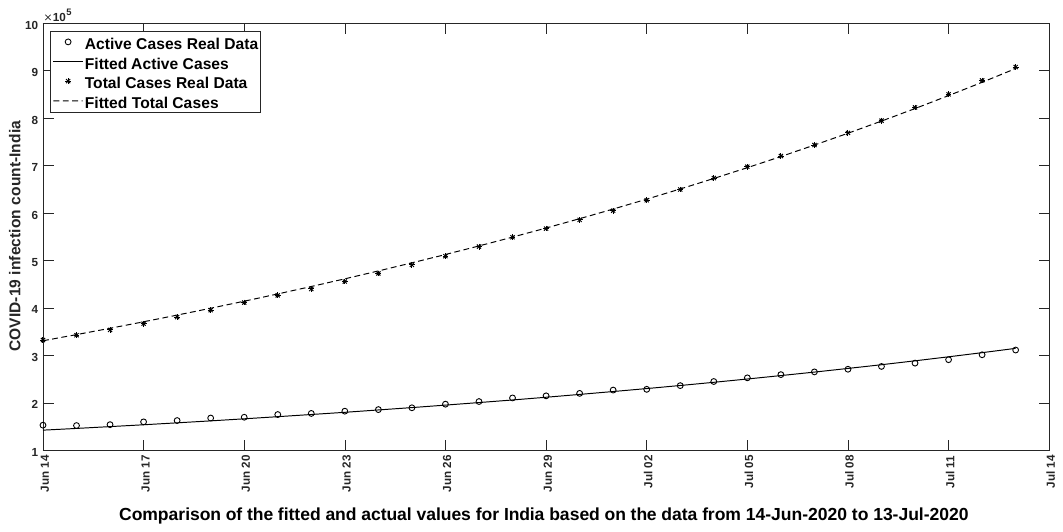

Figure 7

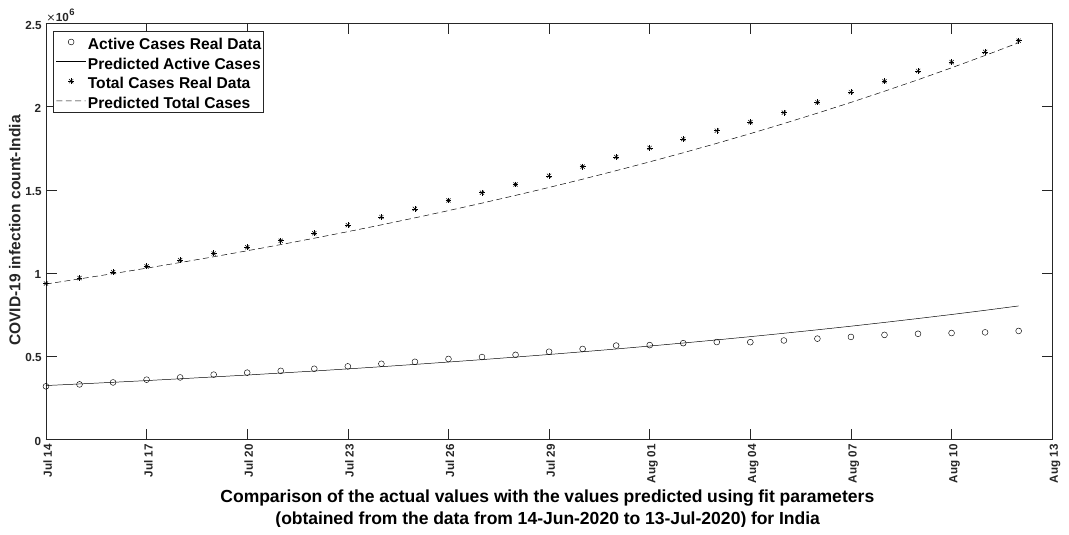

Figure 8

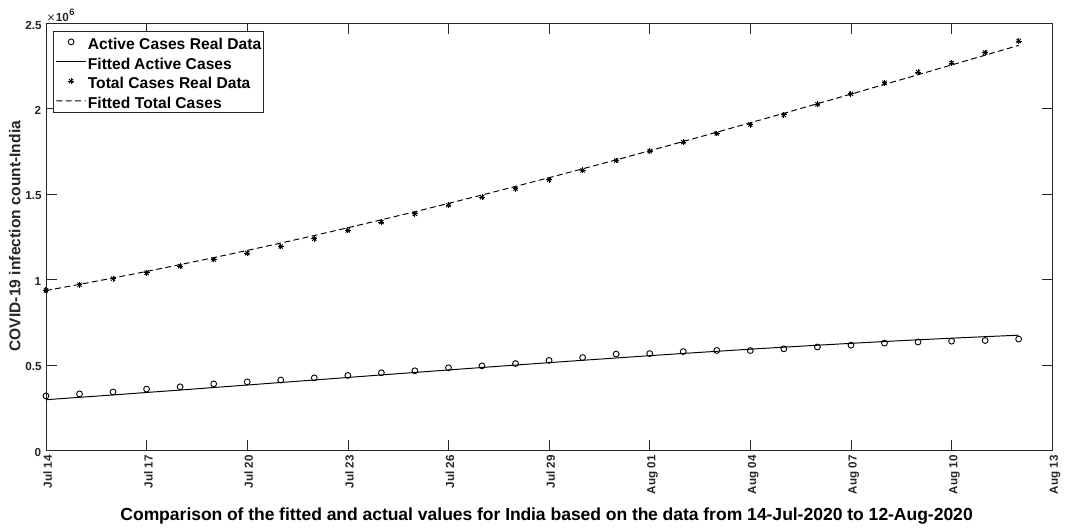

Figure 9

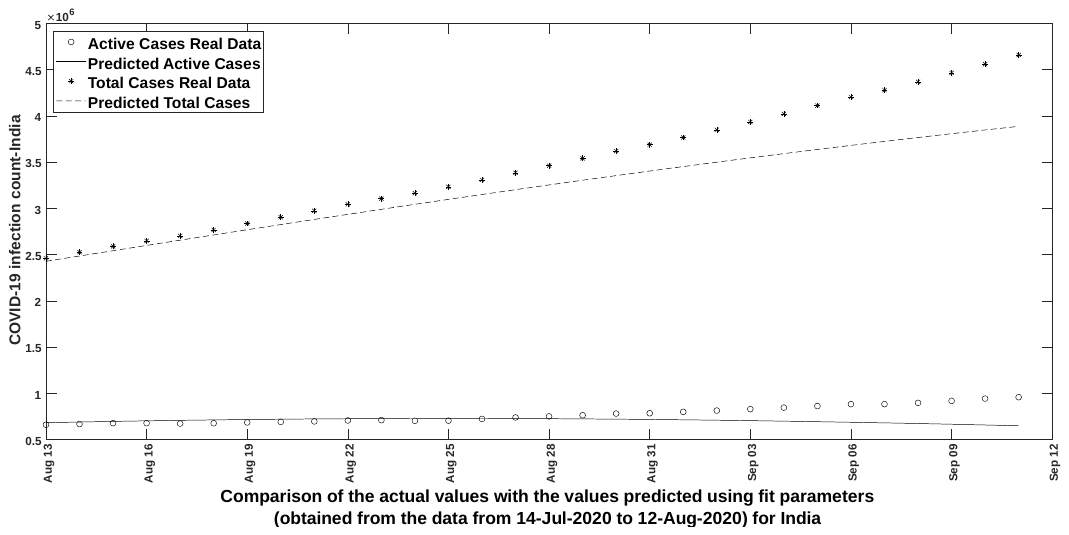

Figure 10

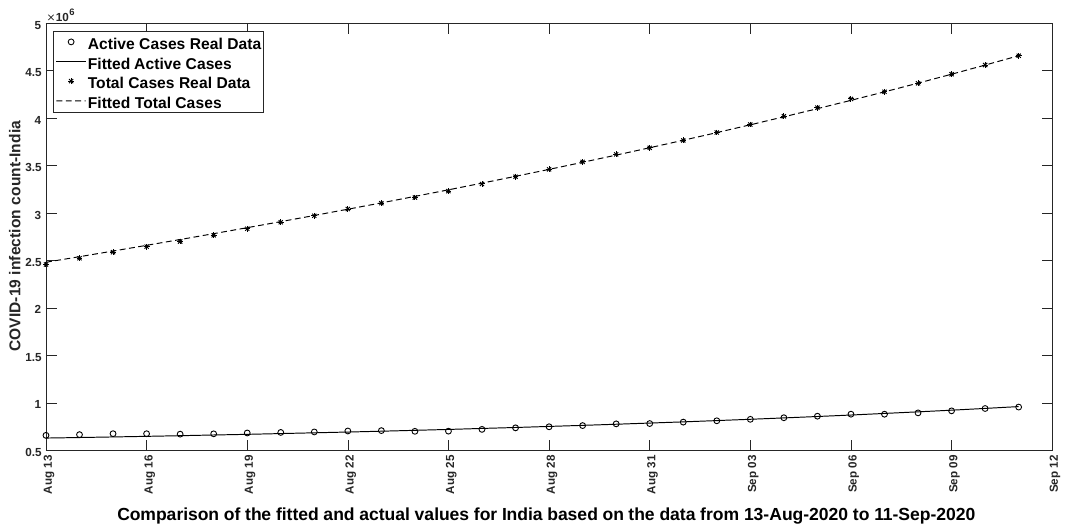

Figure 11

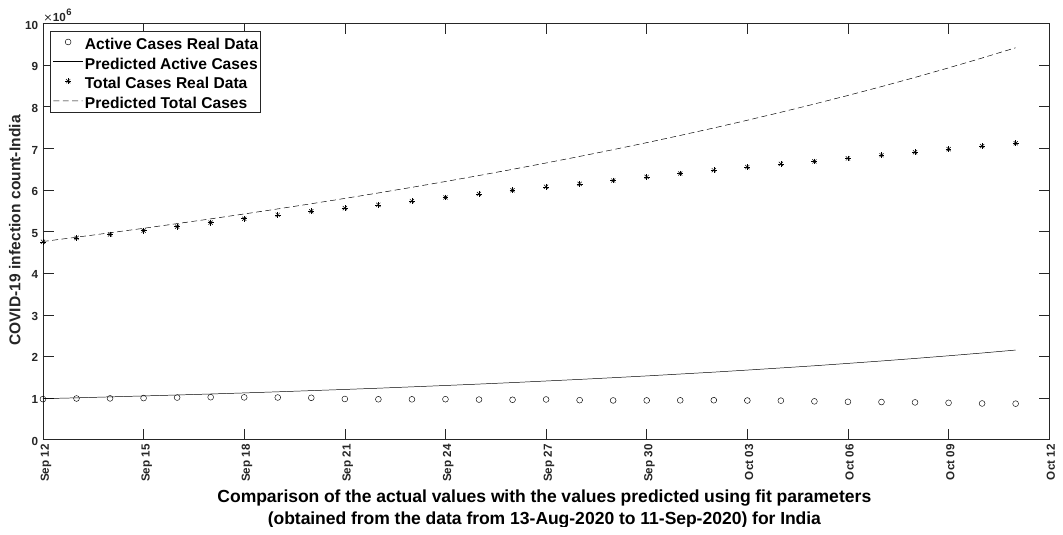

Figure 12

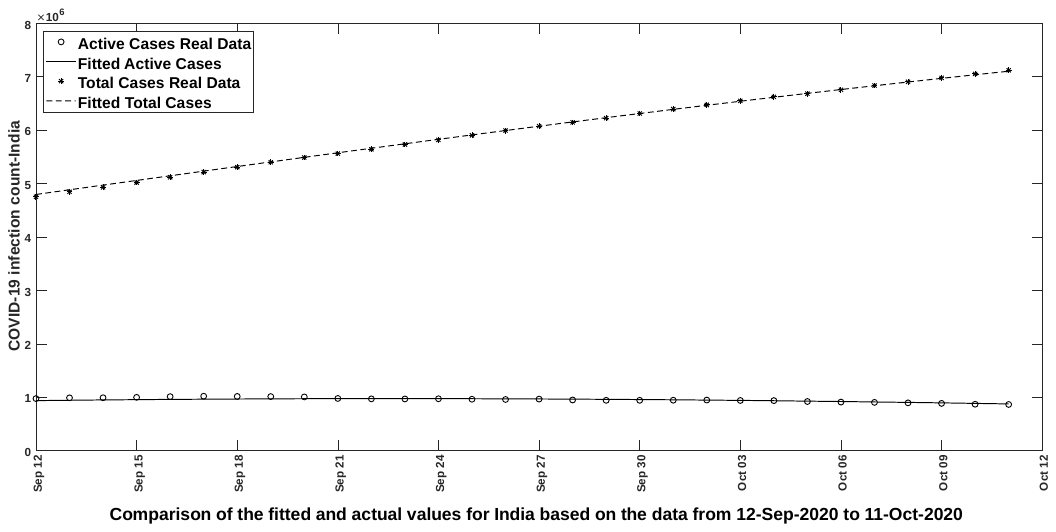

Figure 13

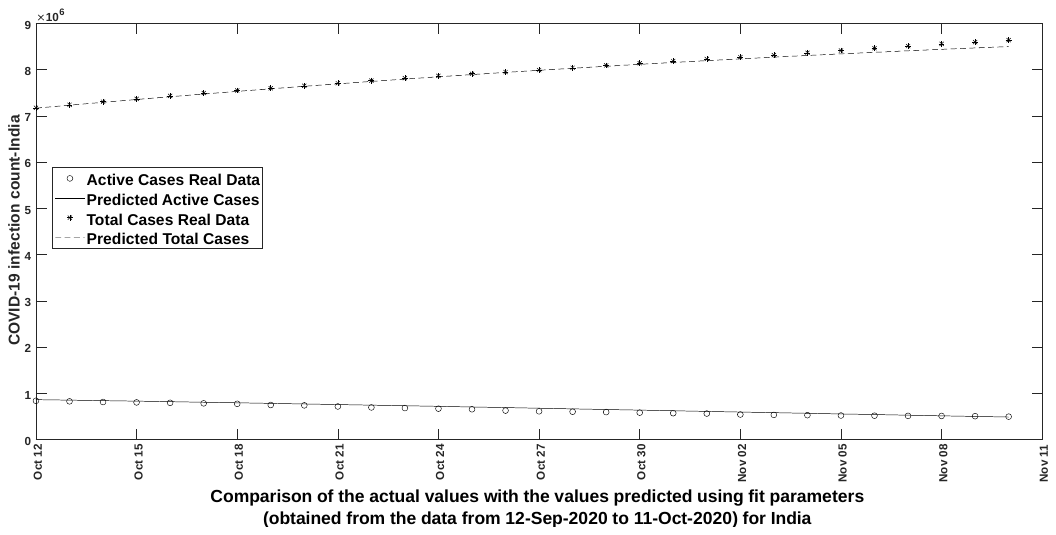

Figure 14

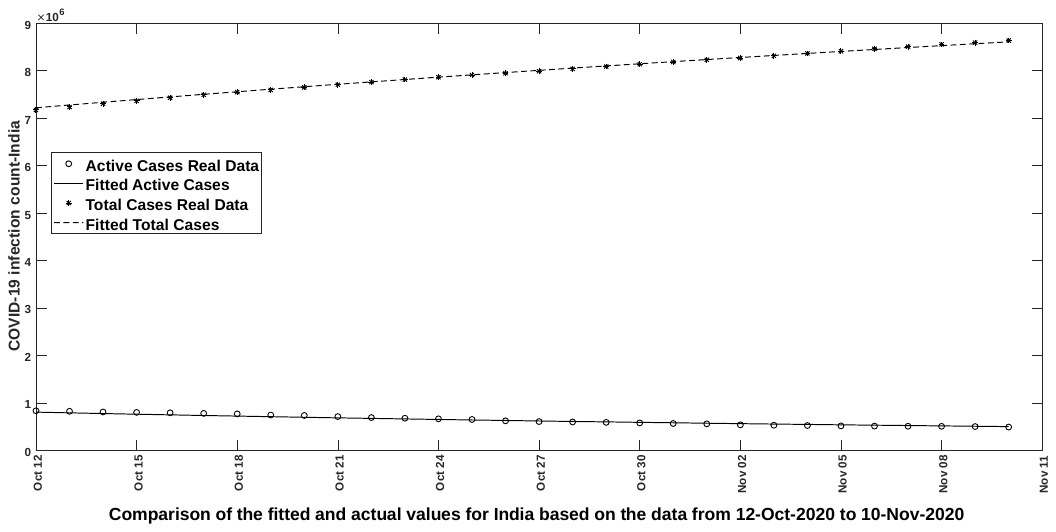

Figure 15

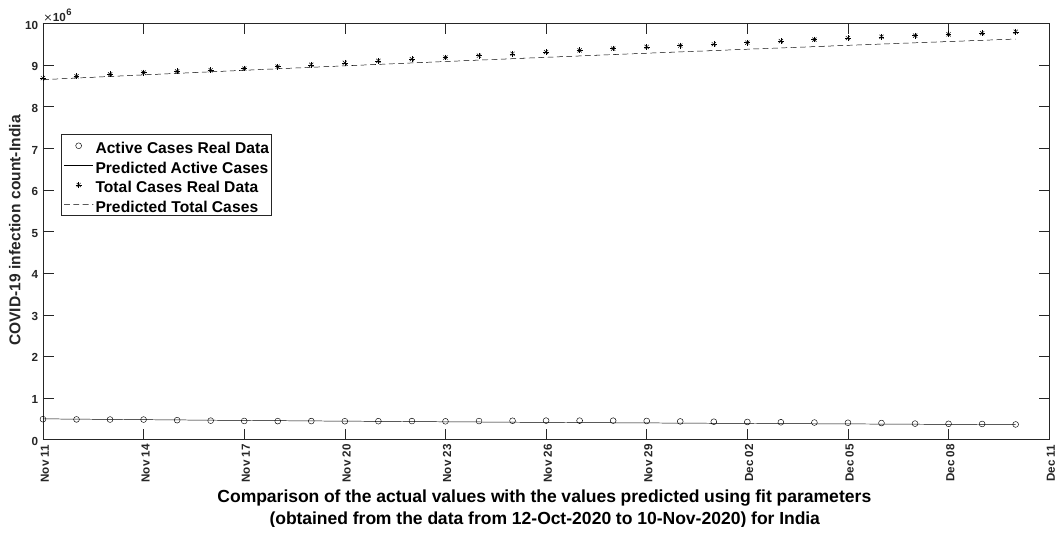

Figure 16

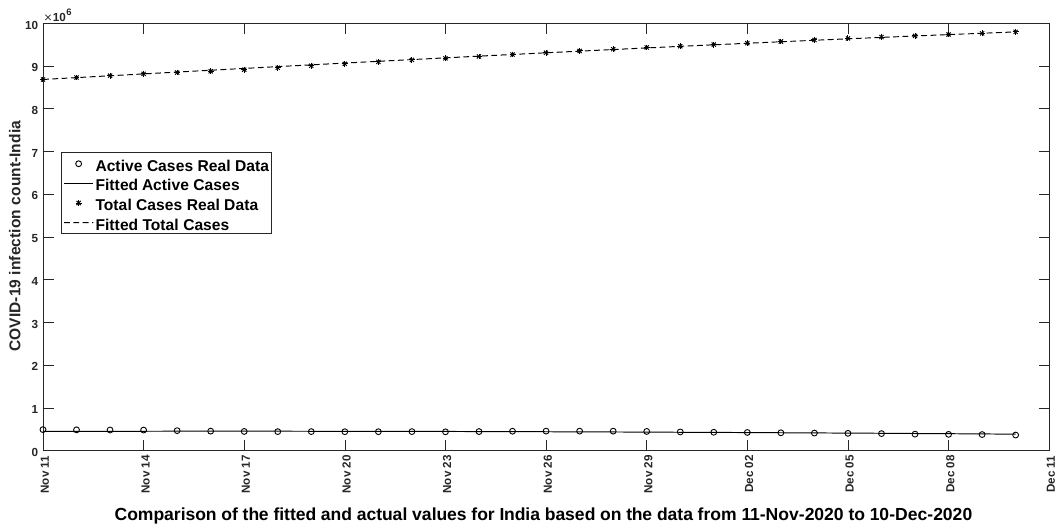

Figure 17

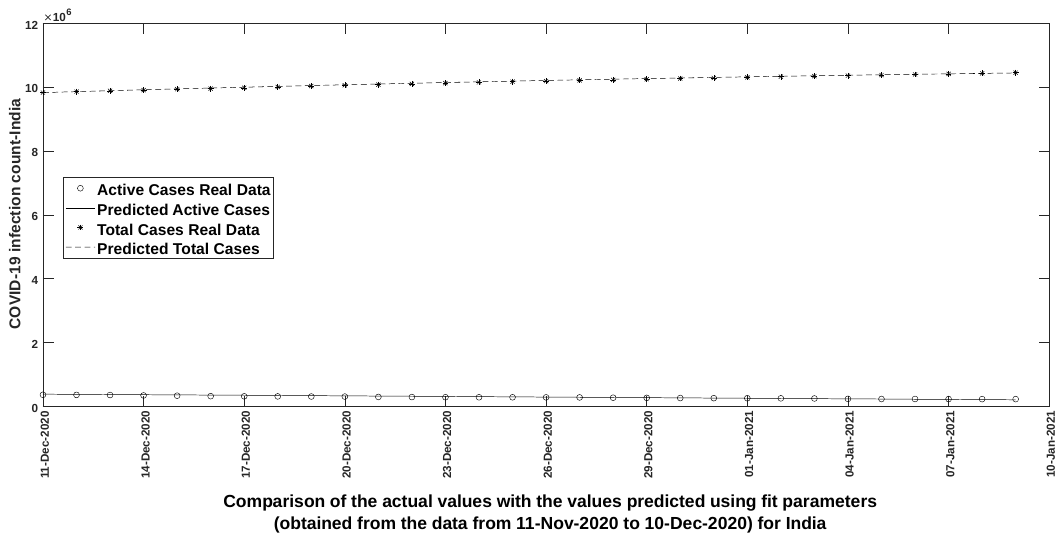

Figure 18

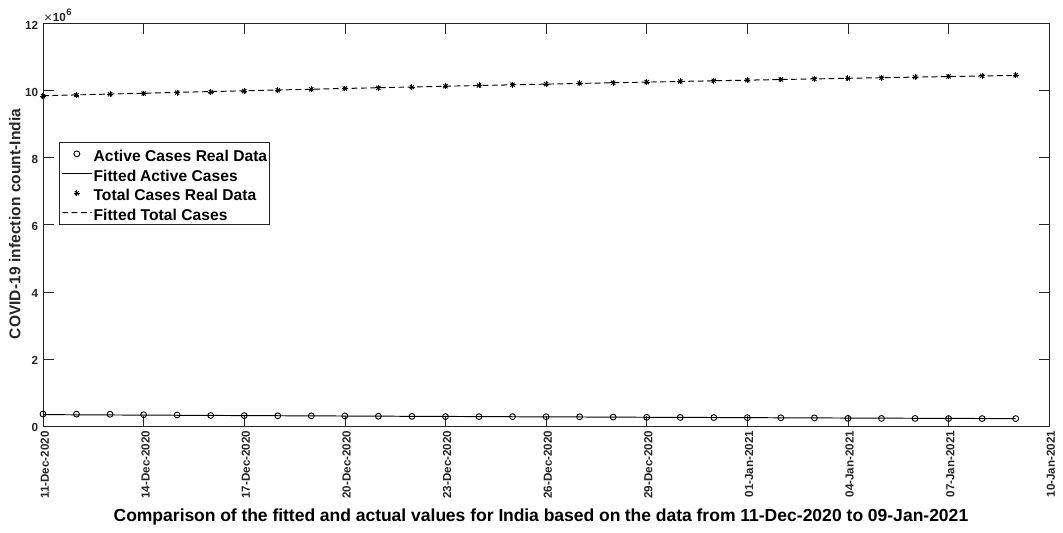

Figure 19

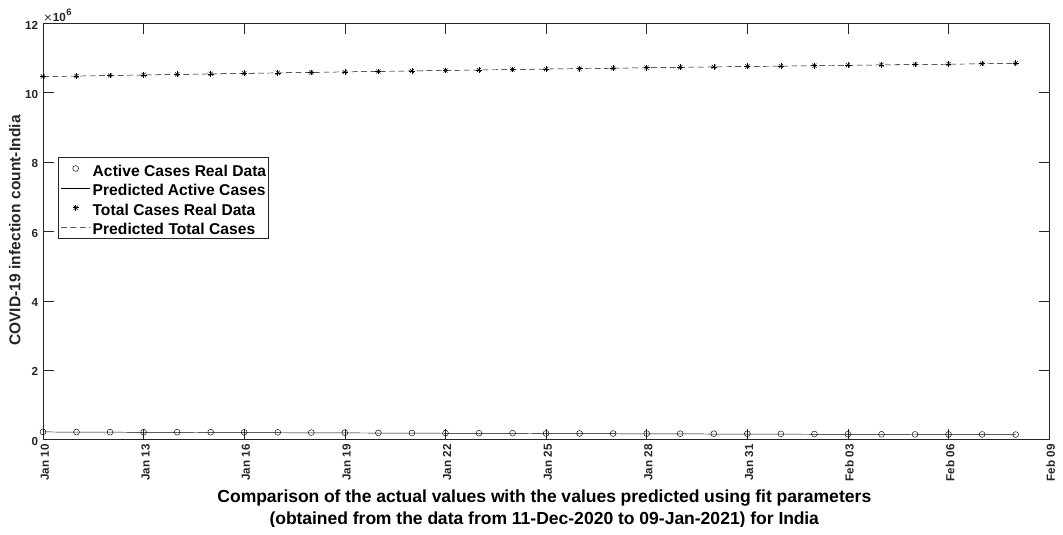

Figure 20

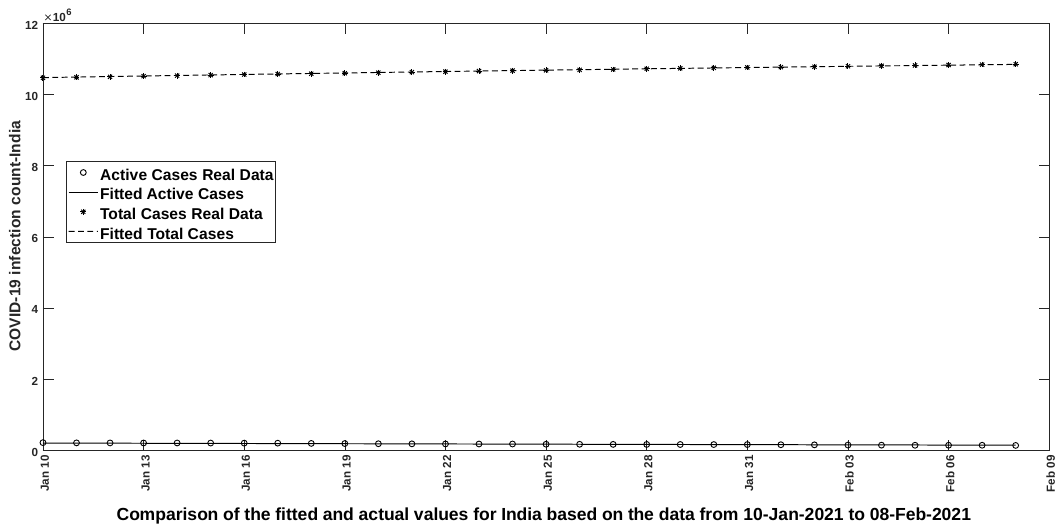

Figure 21

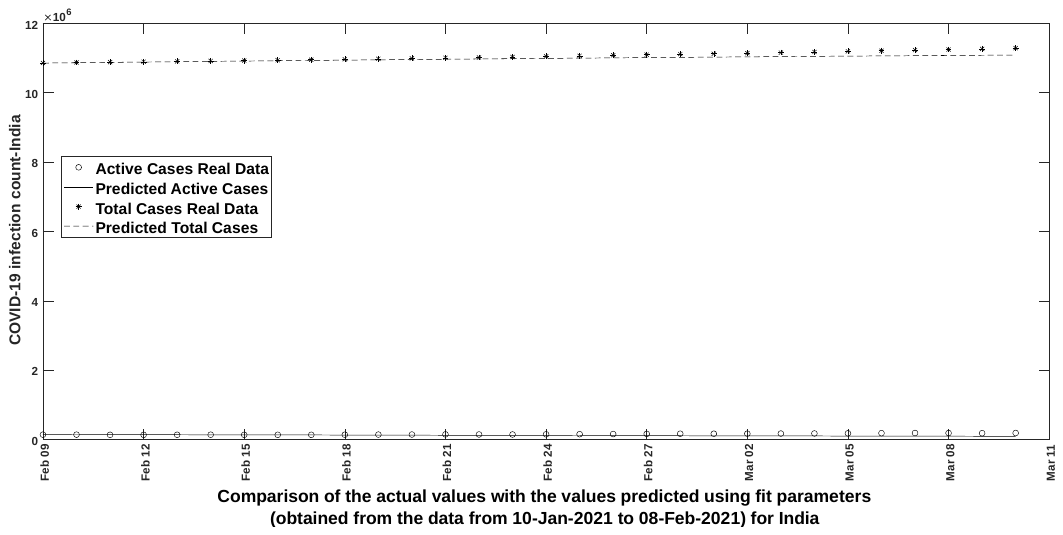

Figure 22

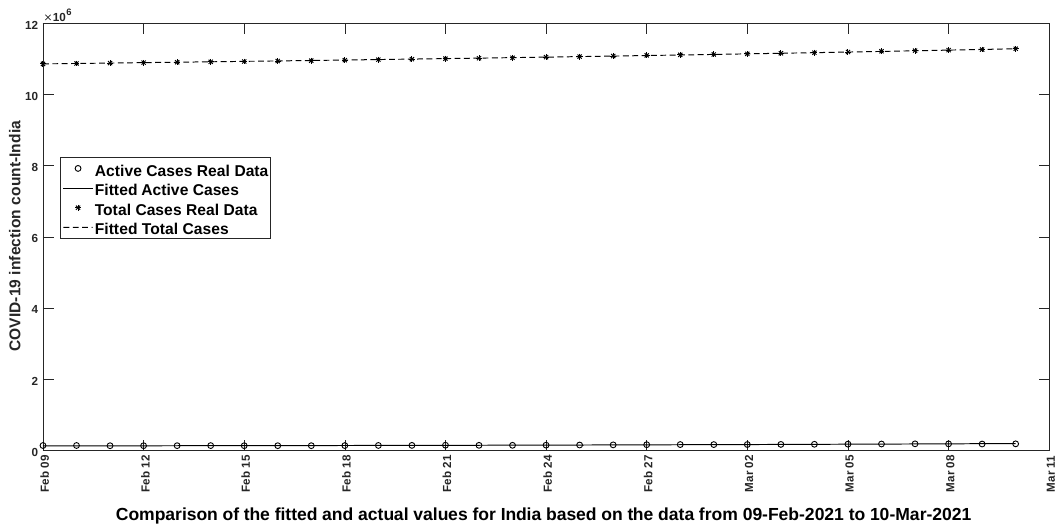

Figure 23

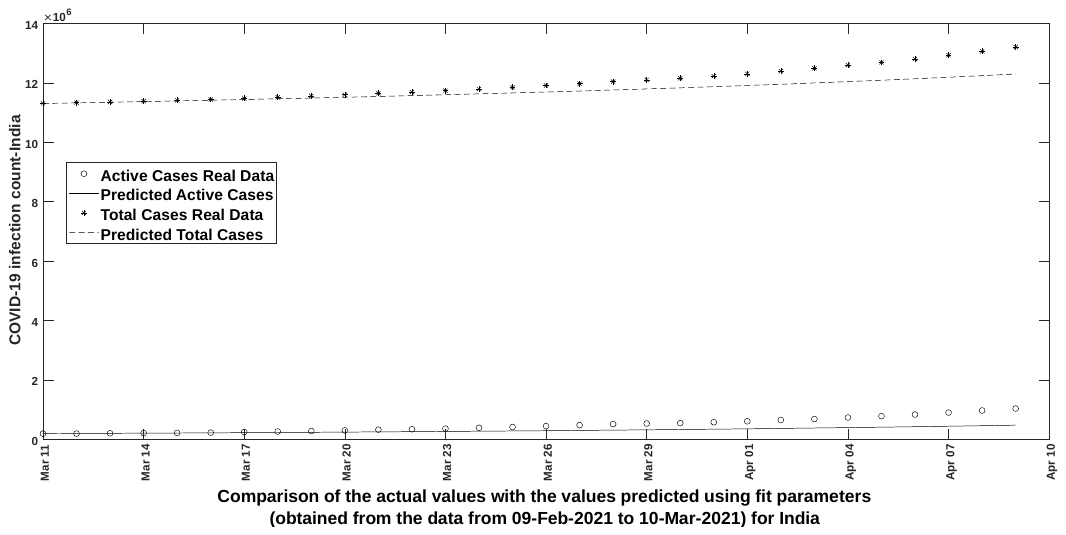

Figure 24

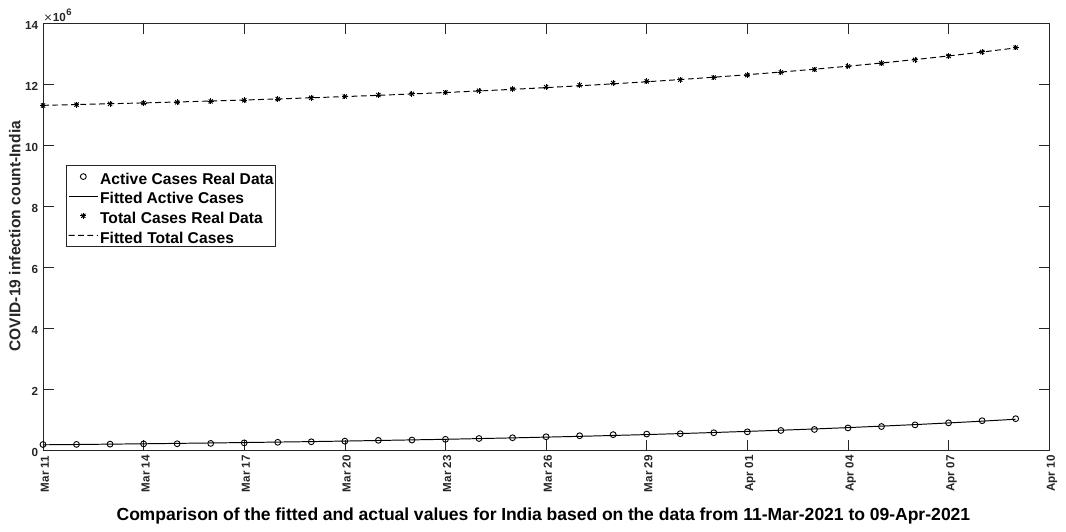

Figure 25

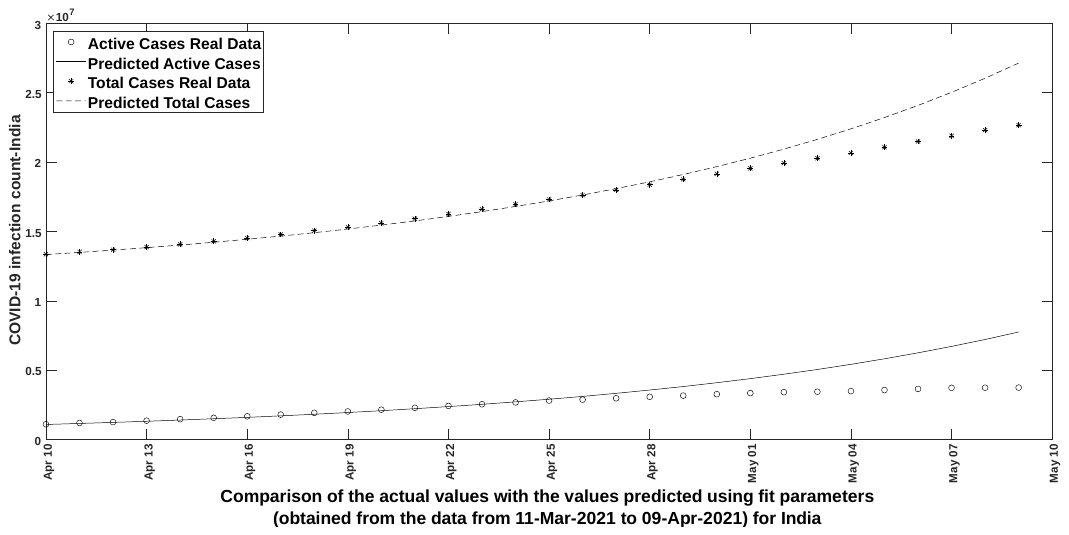

Figure 26

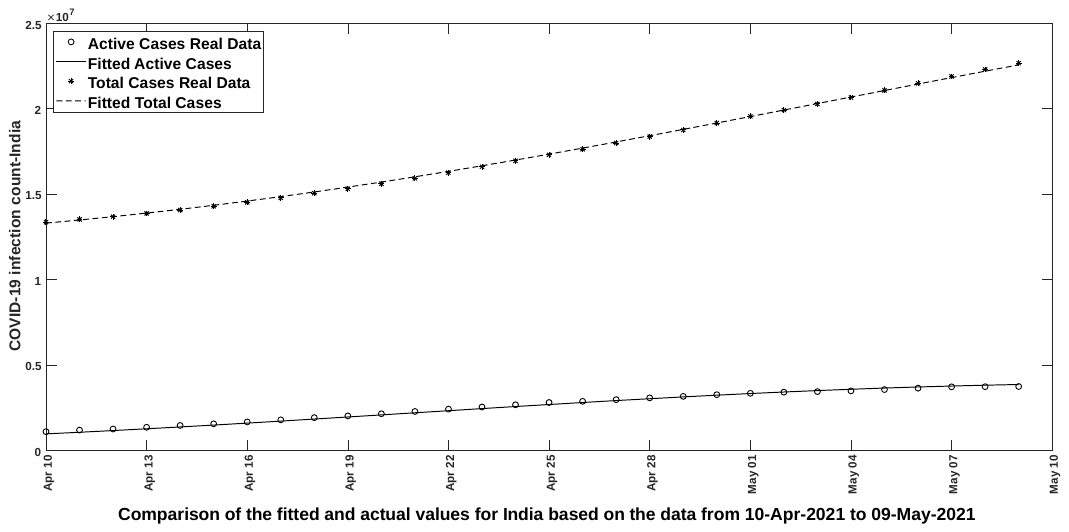

Figure 27

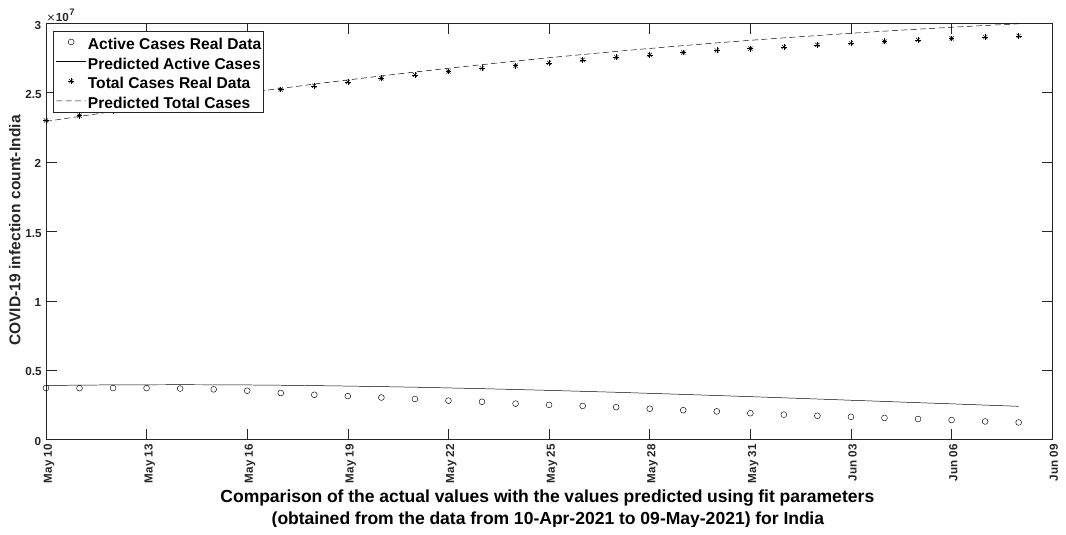

Figure 28

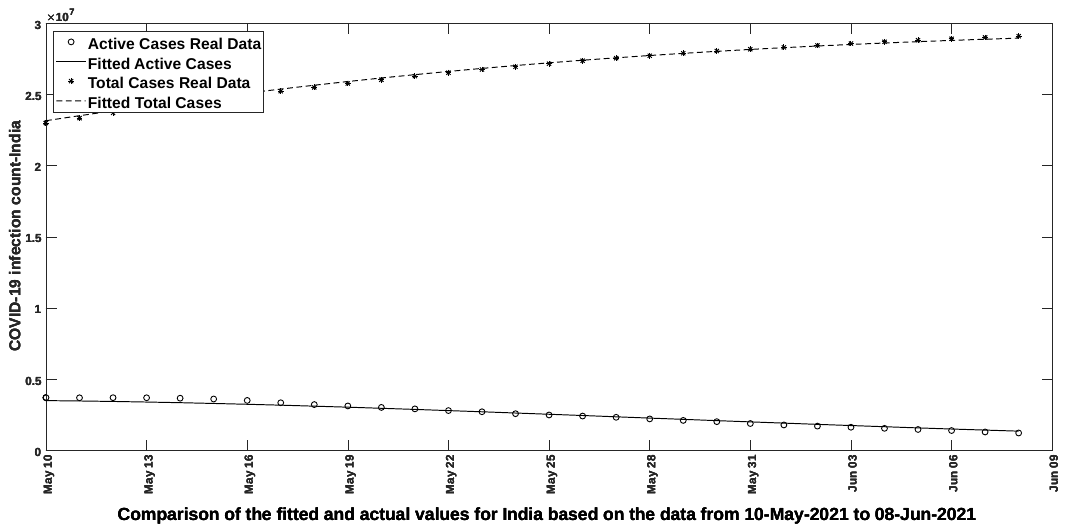

Figure 29

**Appendix II**

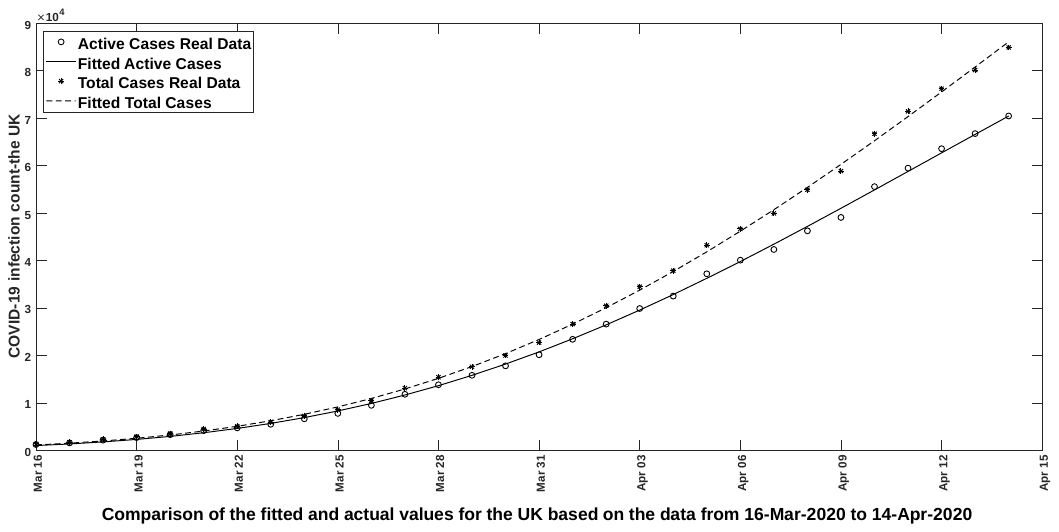

Figure 1

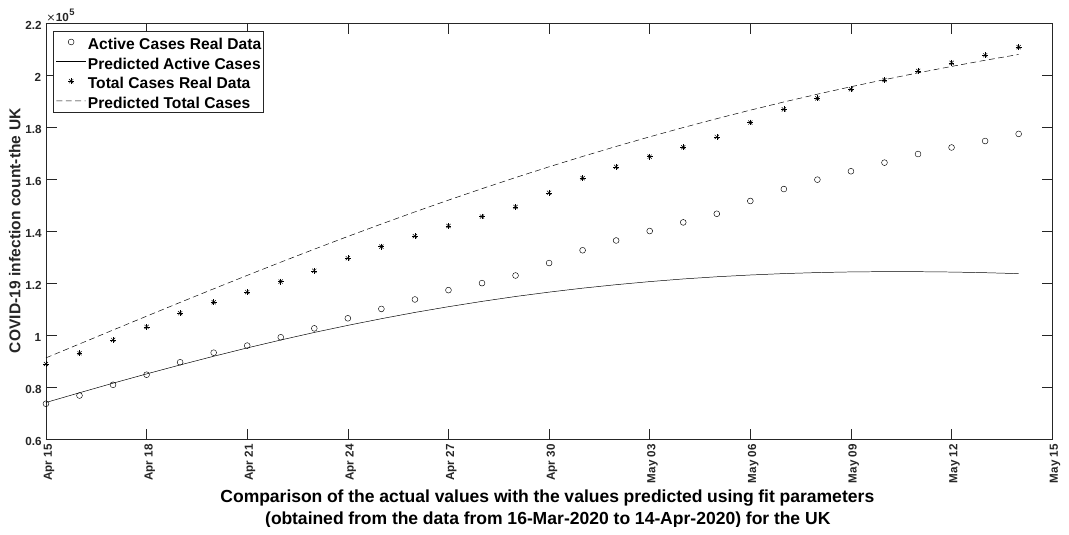

Figure 2

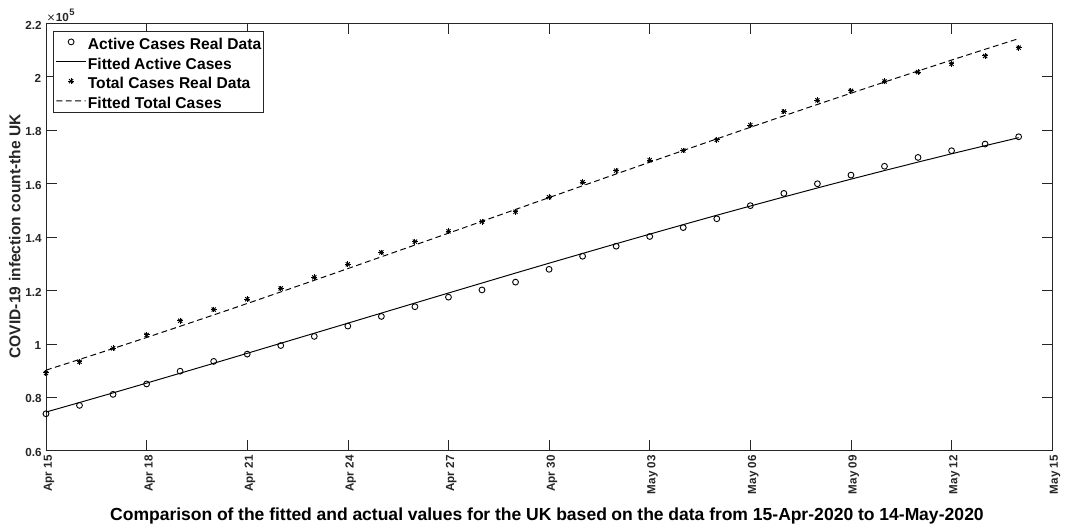

Figure 3

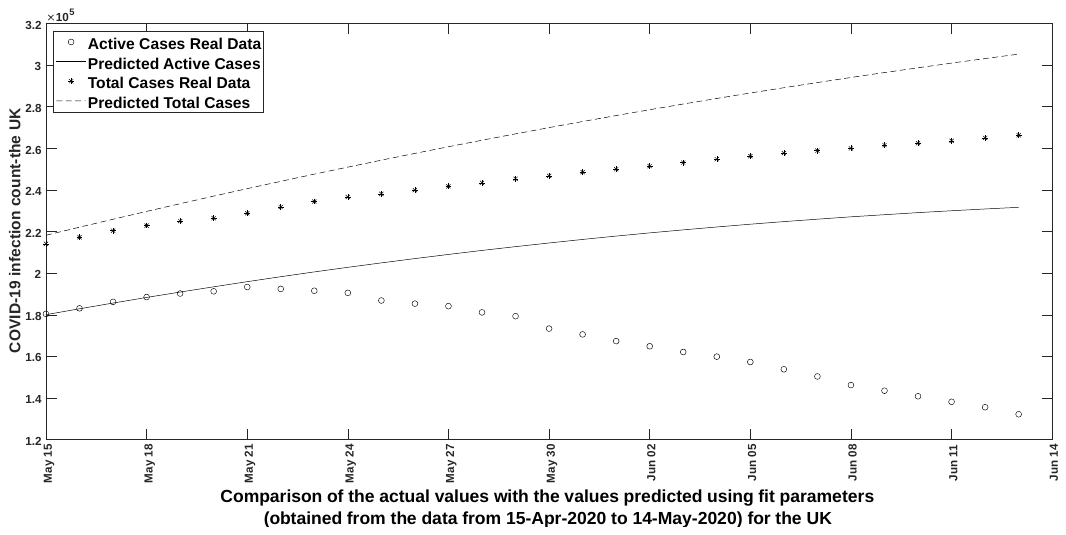

Figure 4

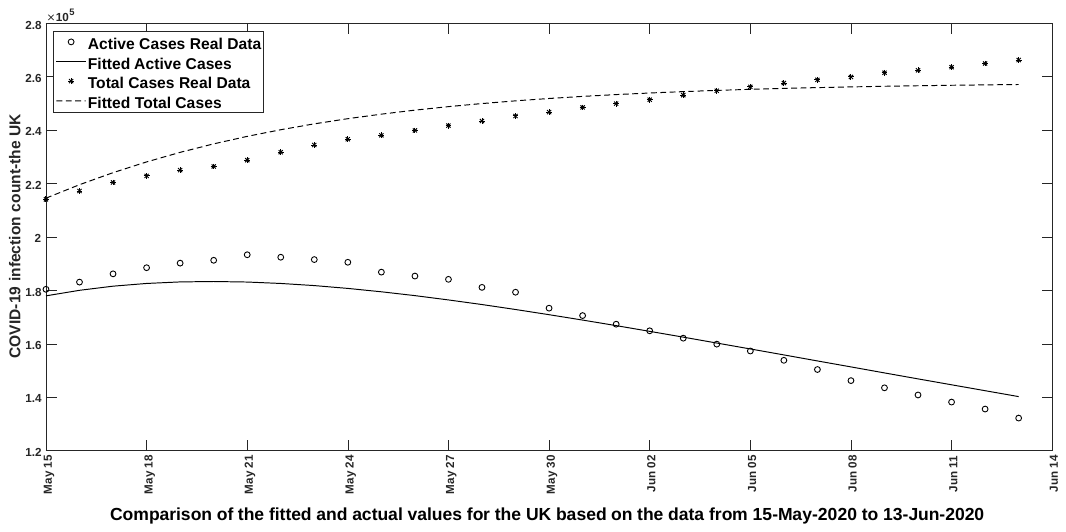

Figure 5

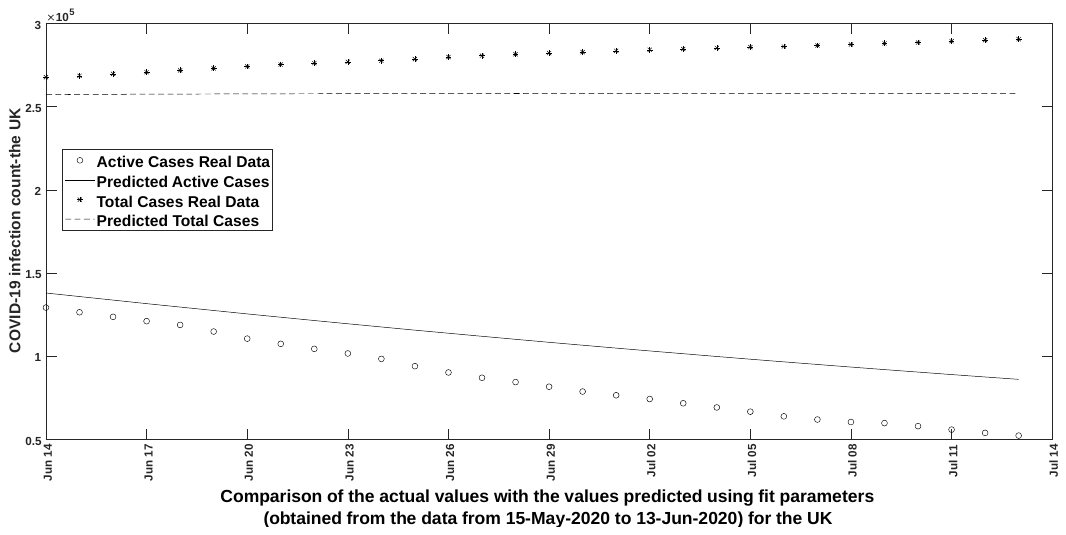

Figure 6

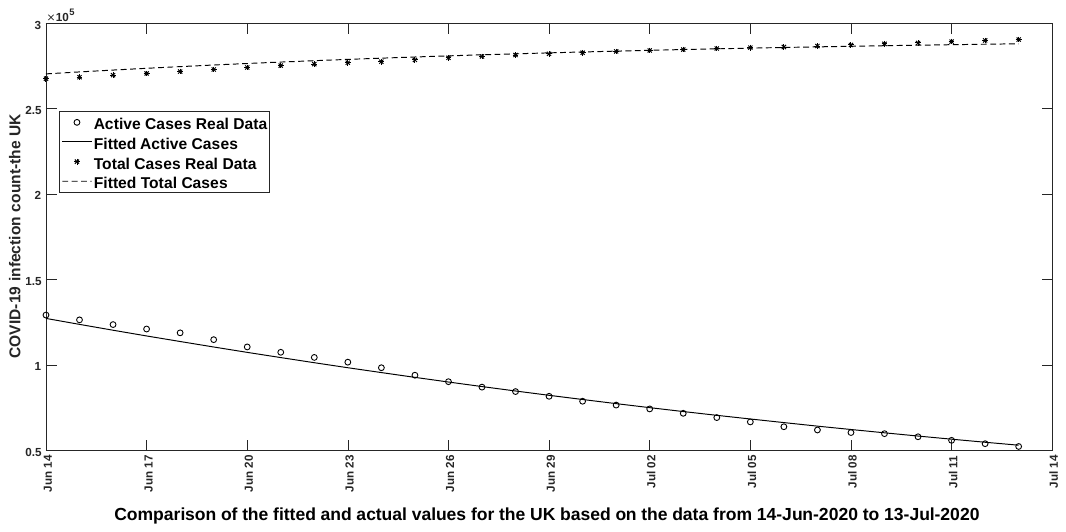

Figure 7

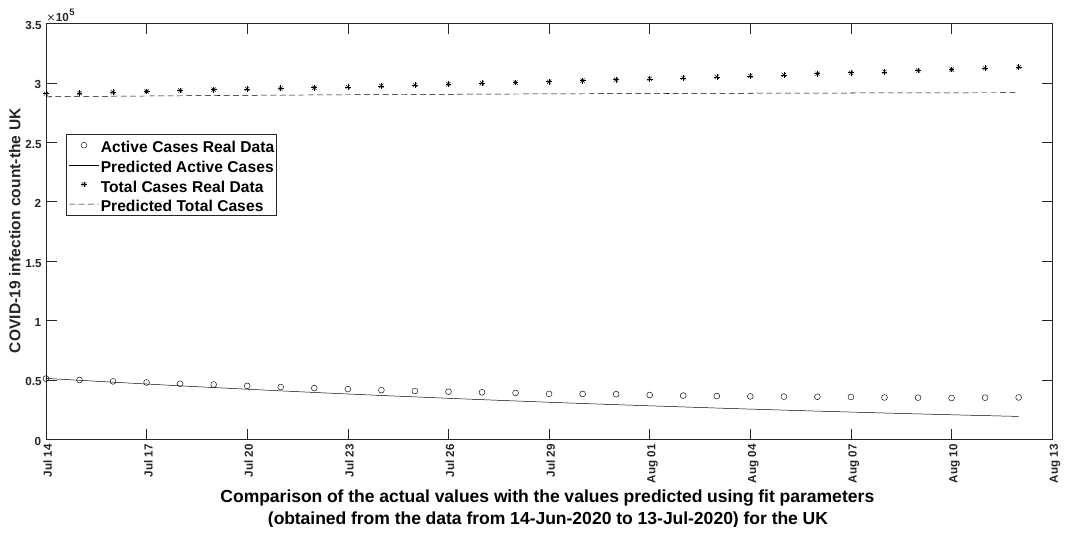

Figure 8

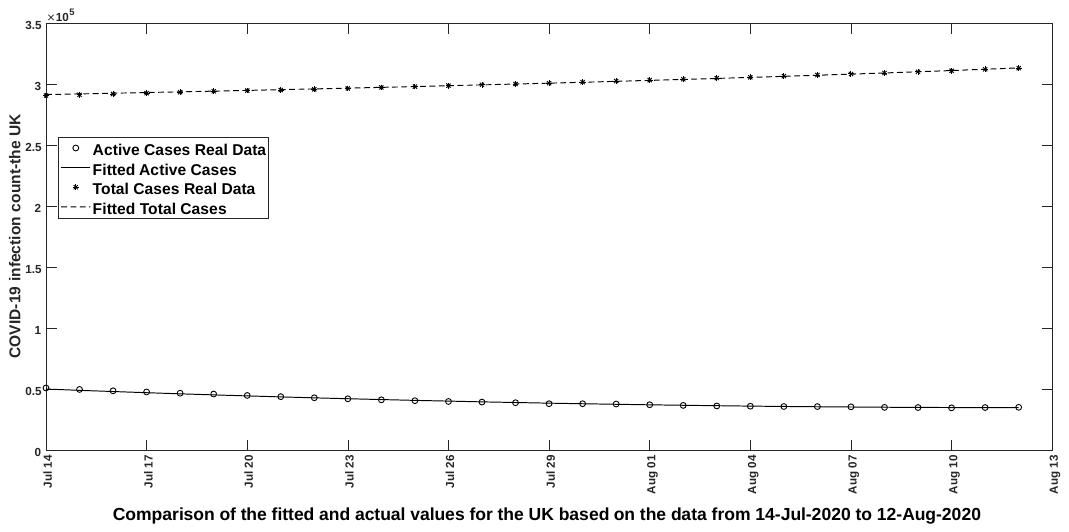

Figure 9

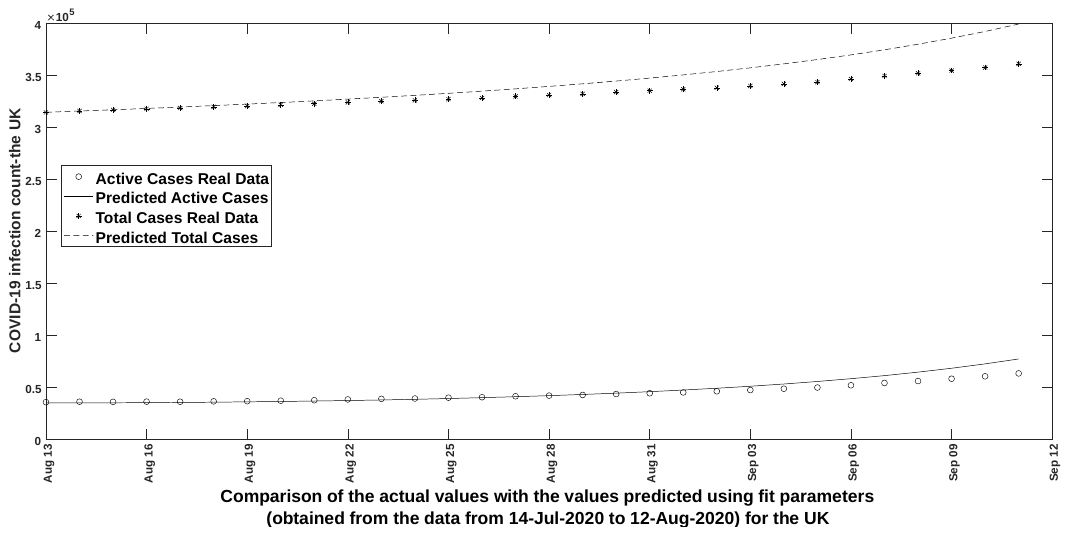

Figure 10

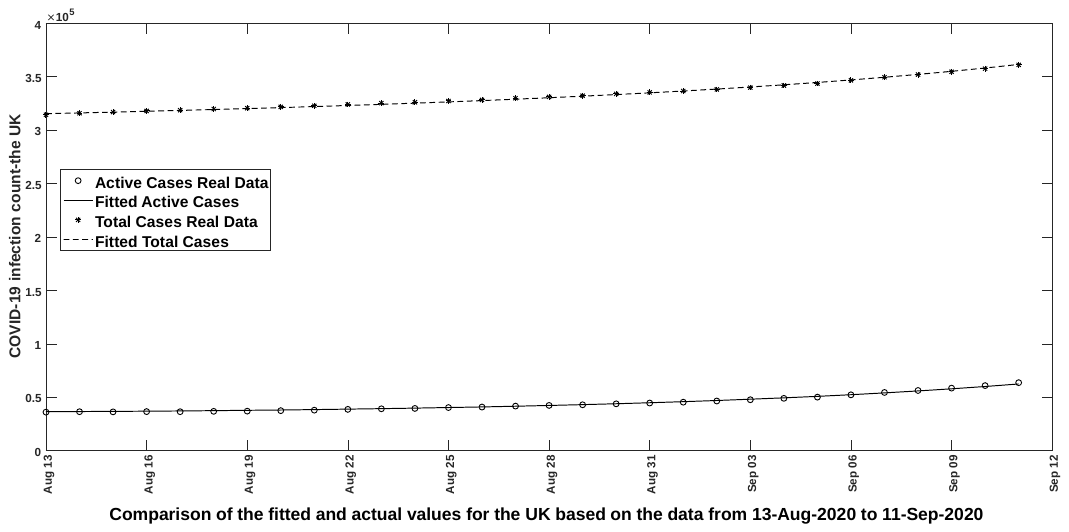

Figure 11

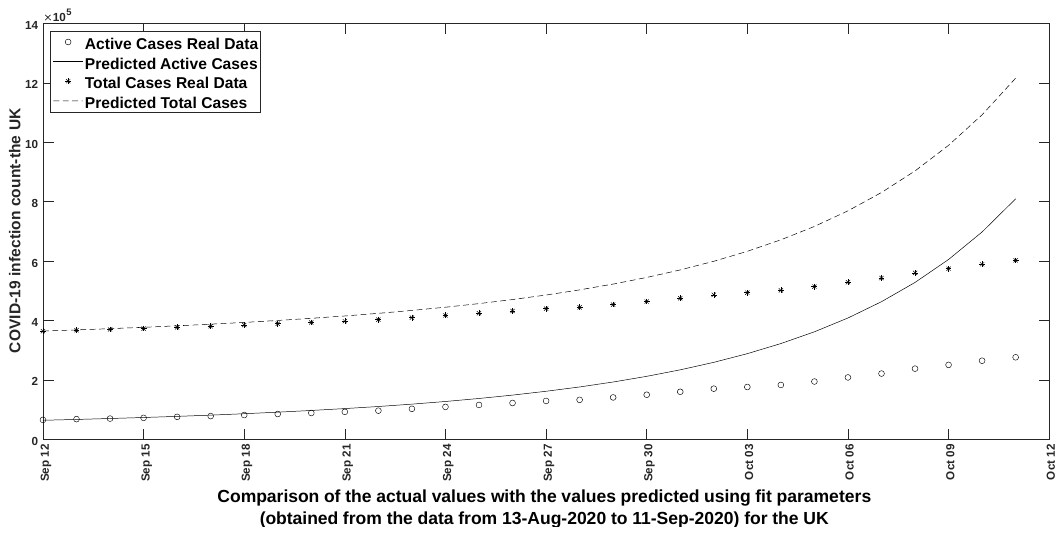

Figure 12

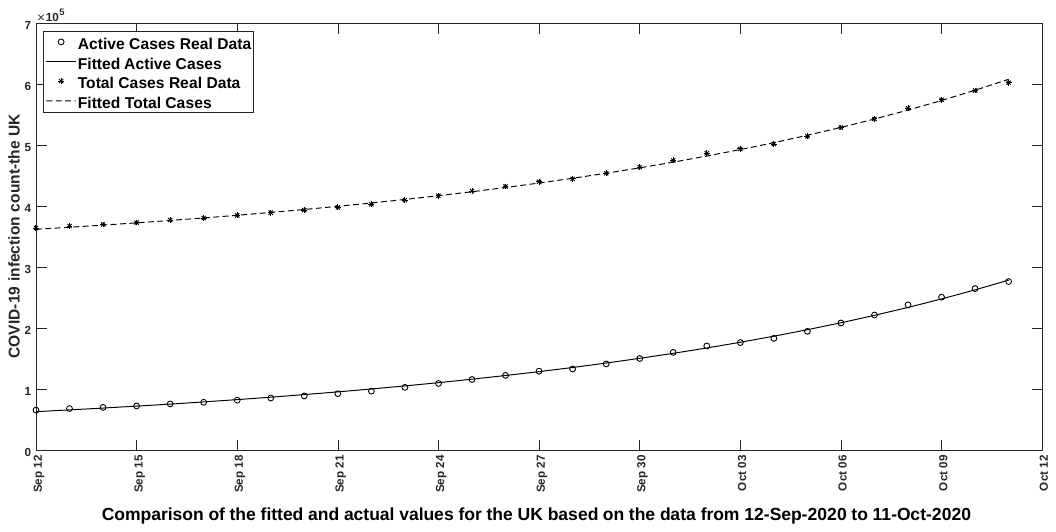

Figure 13

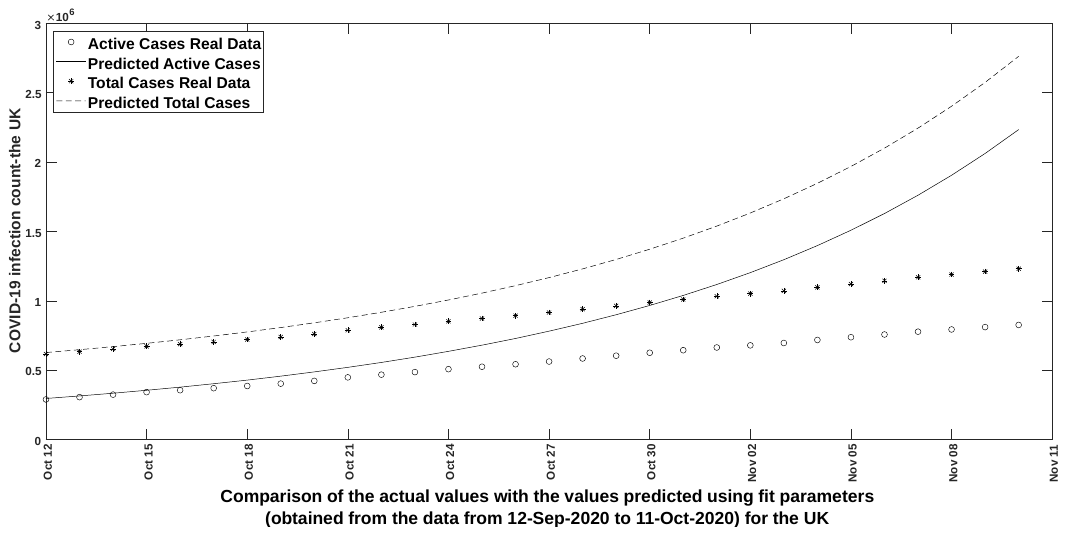

Figure 14

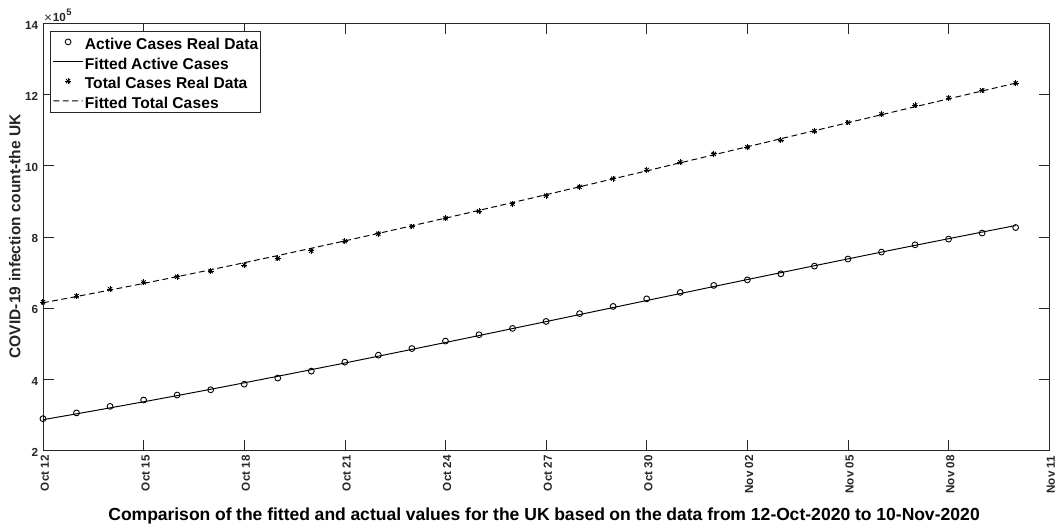

Figure 15

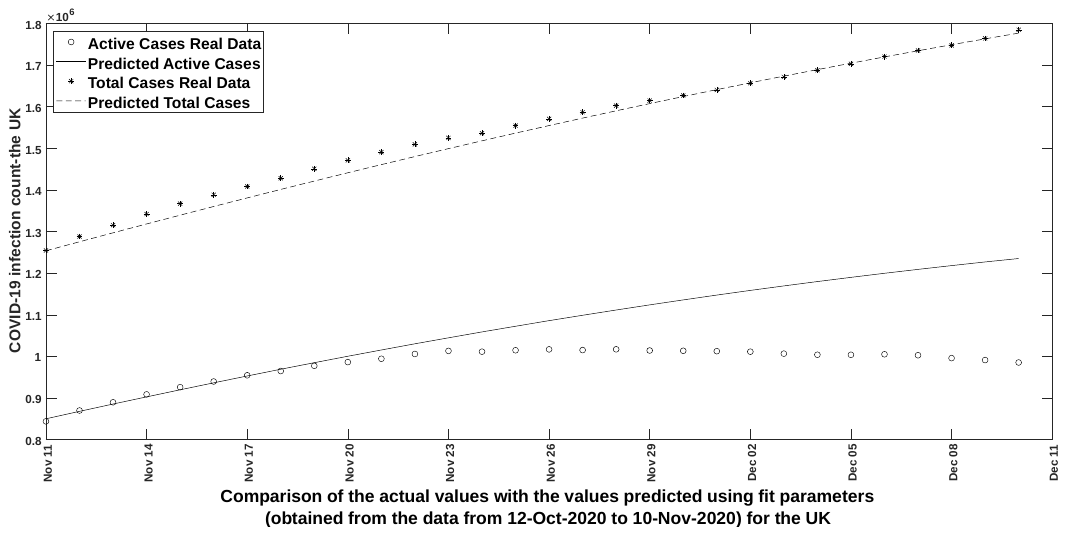

Figure 16

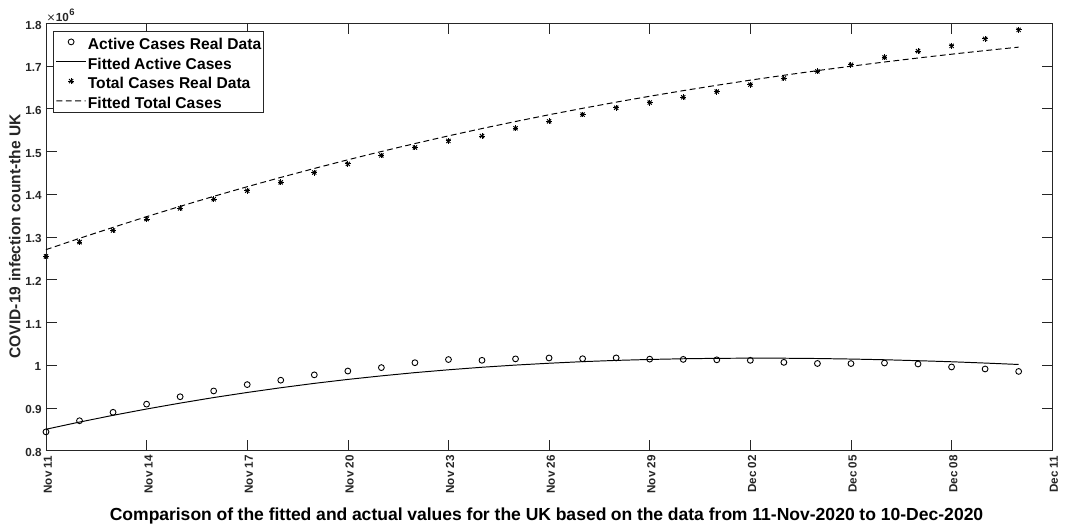

Figure 17

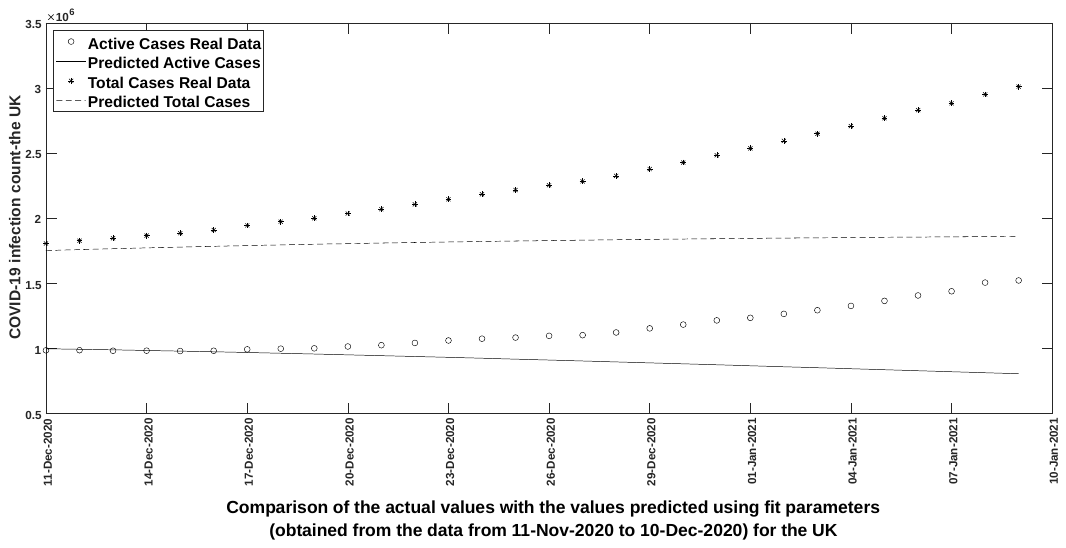

Figure 18

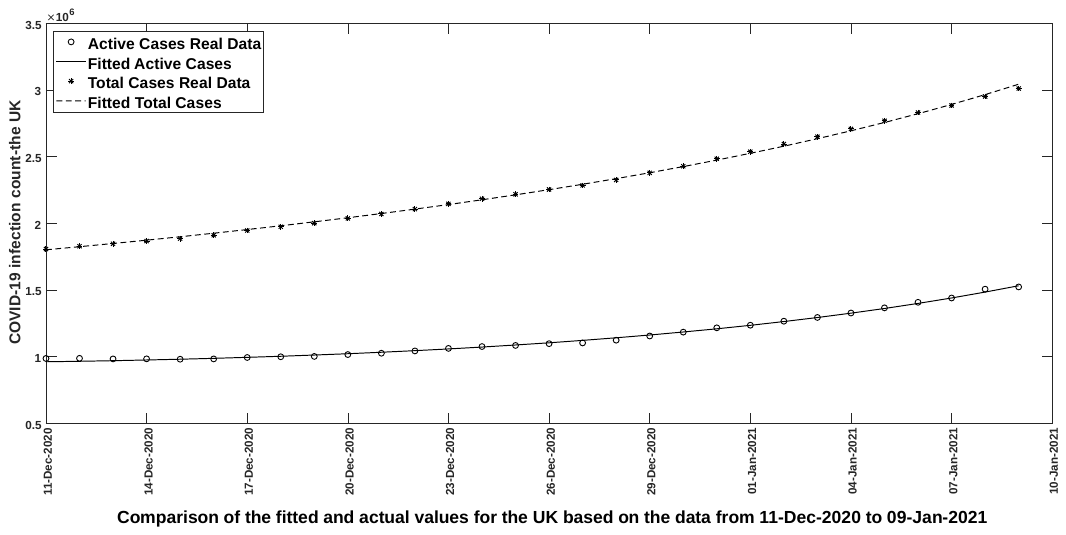

Figure 19

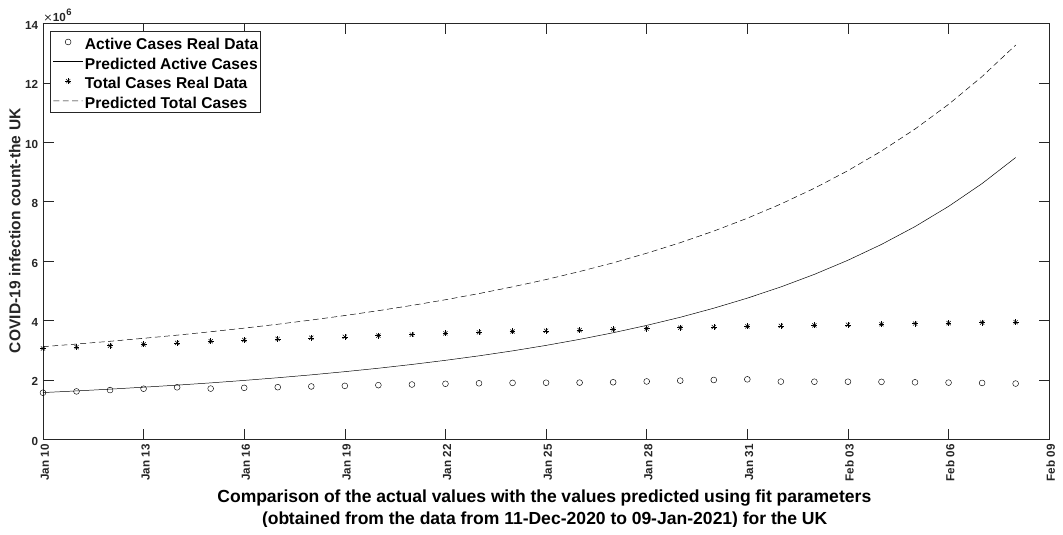

Figure 20

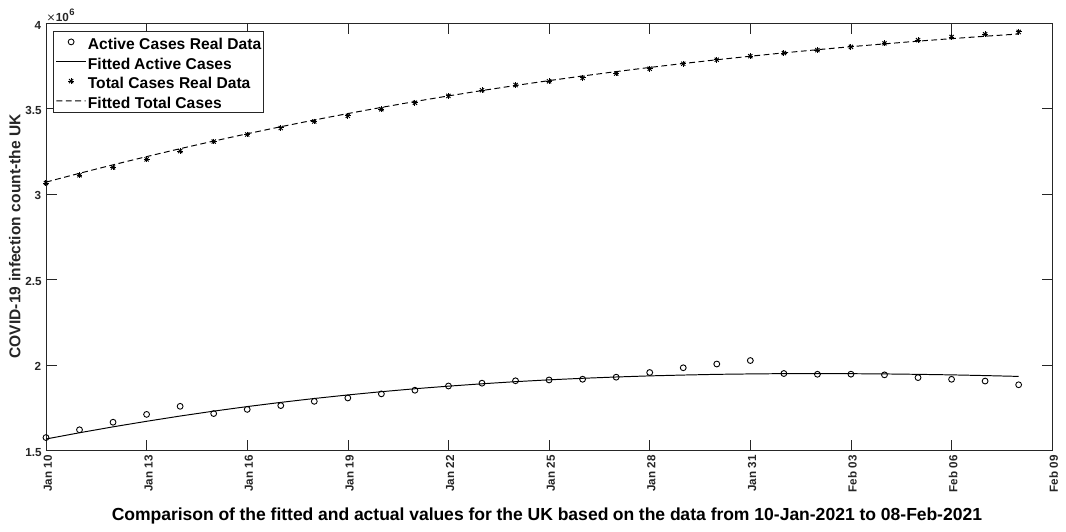

Figure 21

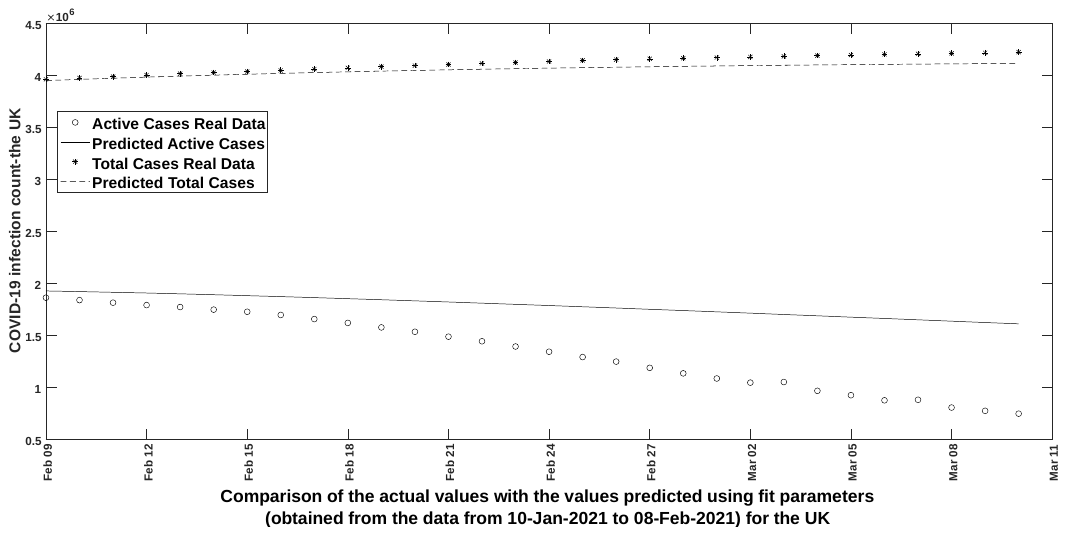

Figure 22

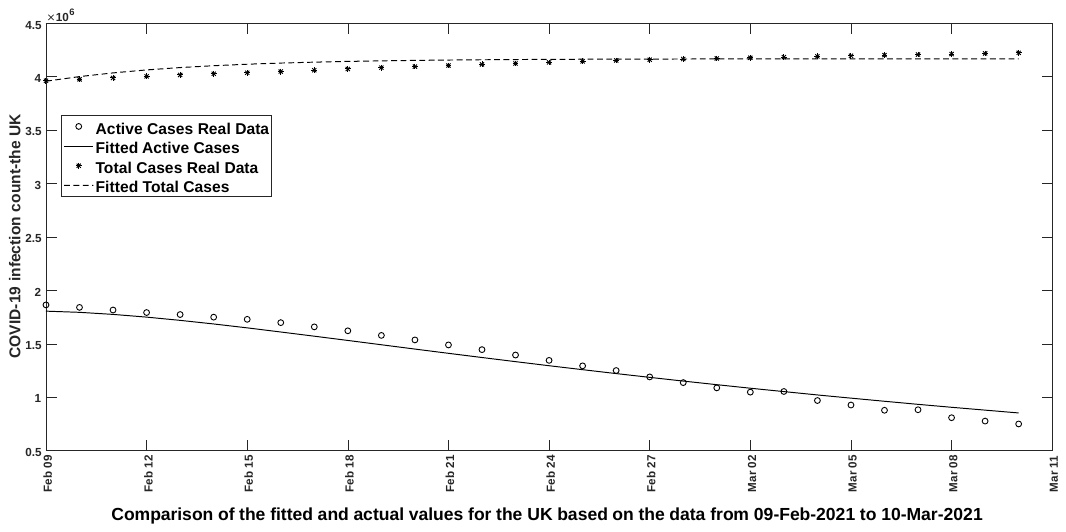

Figure 23

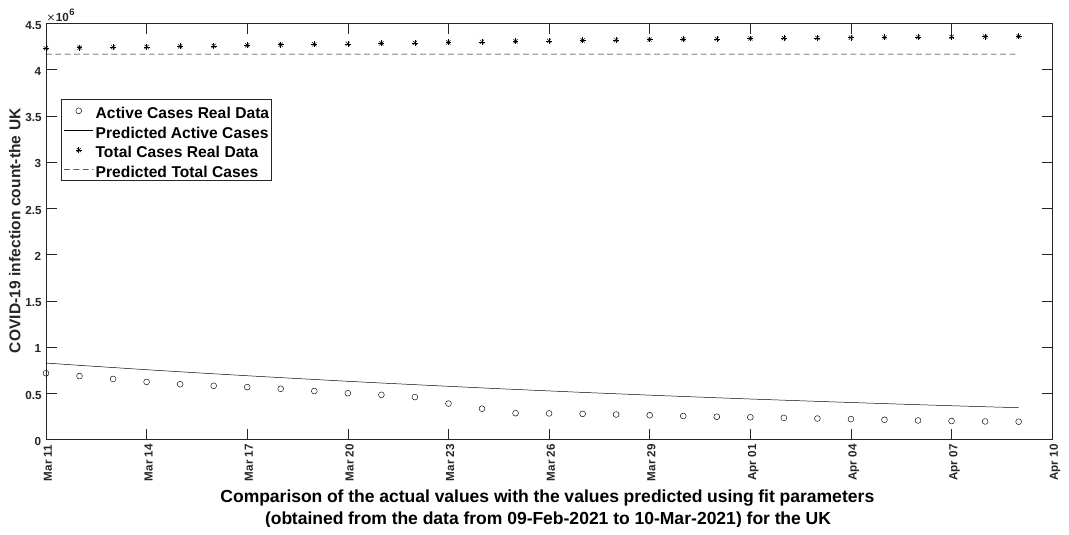

Figure 24

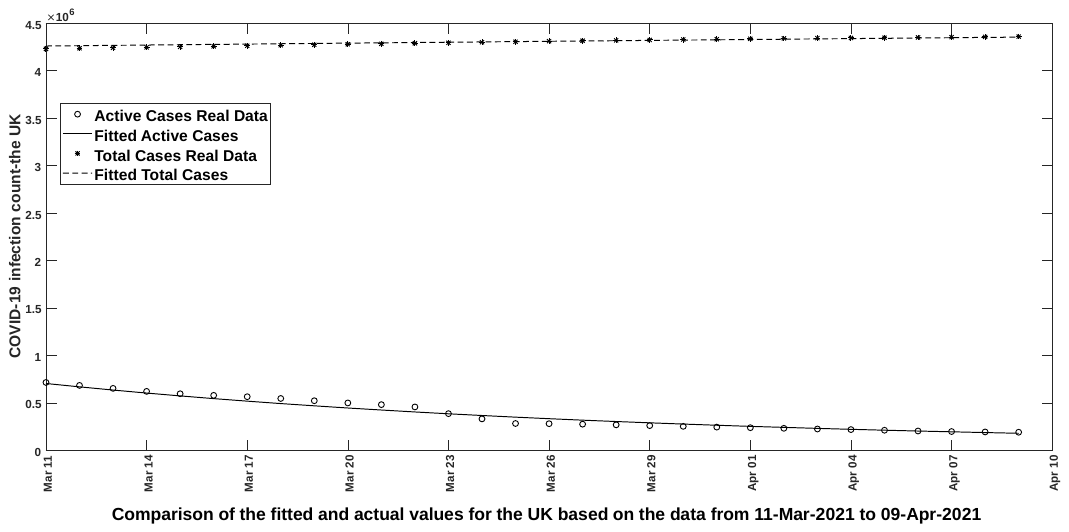

Figure 25

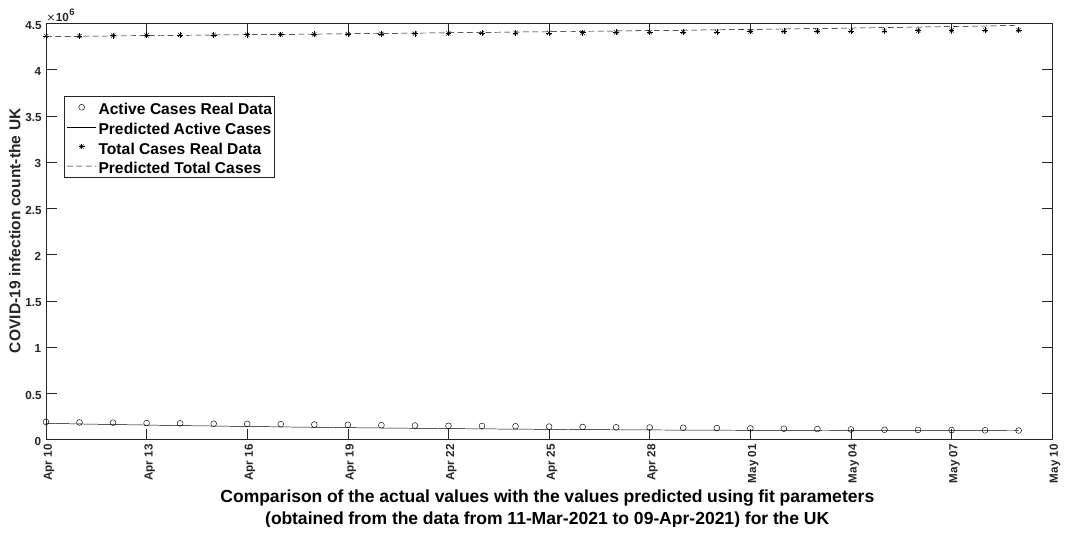

Figure 26

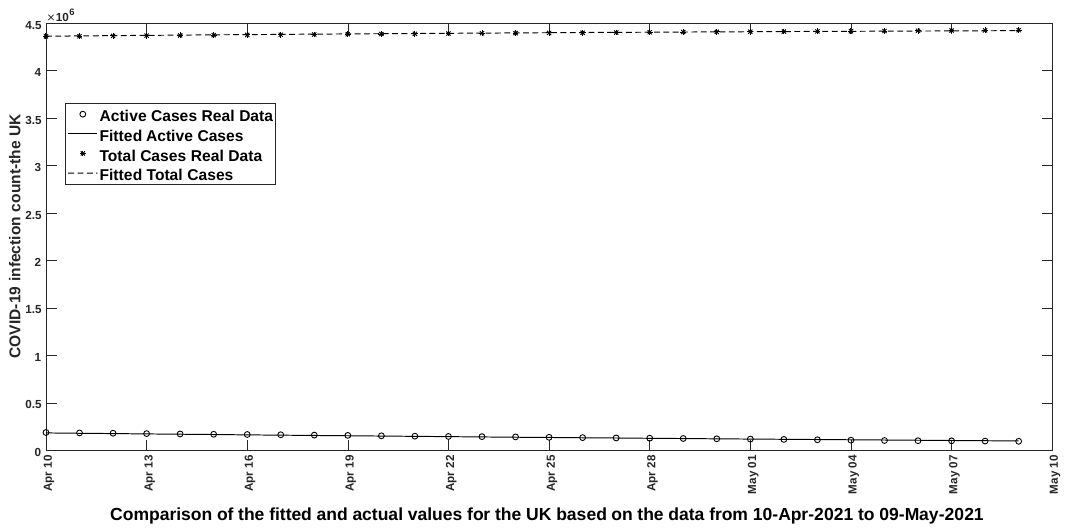

Figure 27

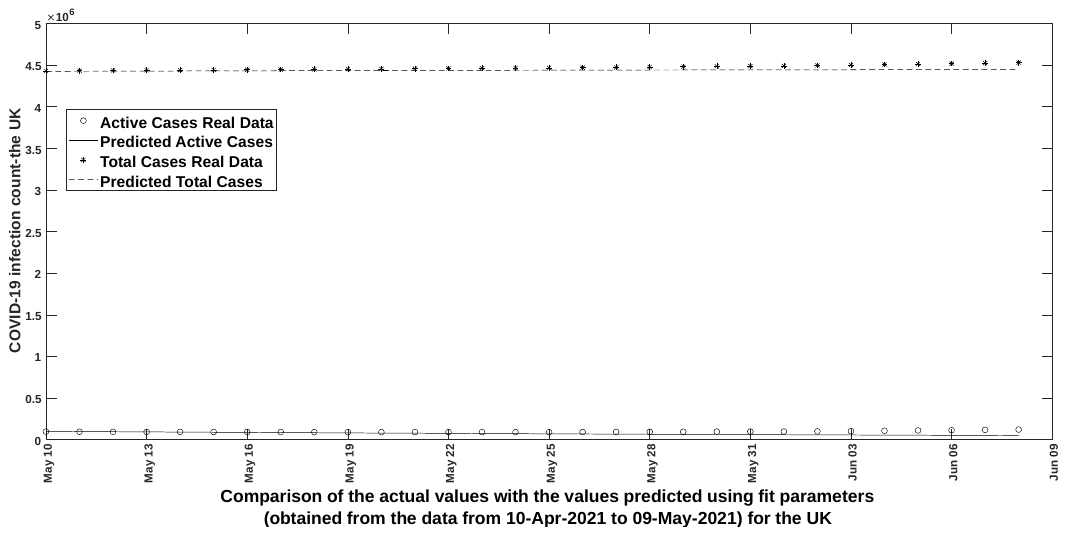

Figure 28

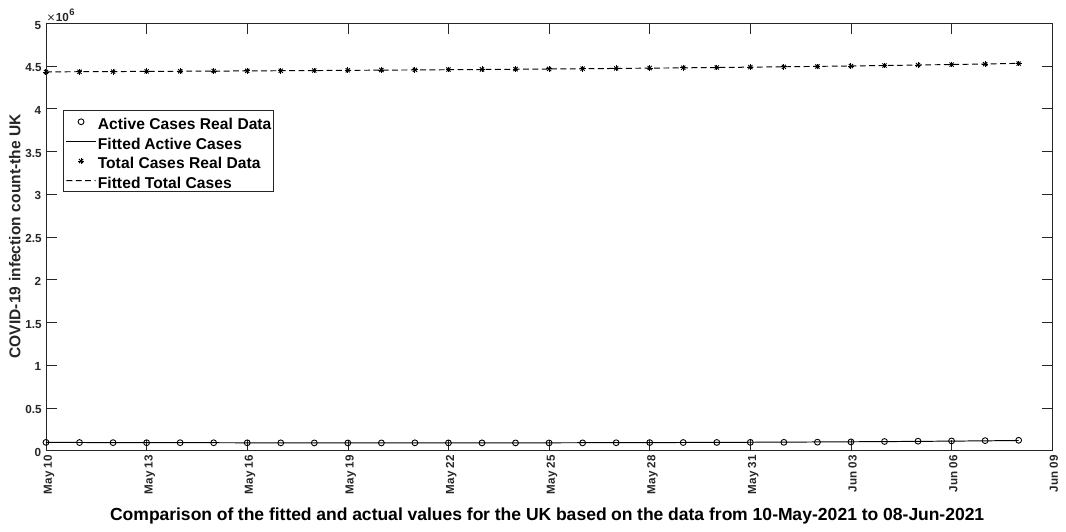

Figure 29
